# Age-dependent impact of the major common genetic risk factor for COVID-19 on severity and mortality

**DOI:** 10.1101/2021.03.07.21252875

**Authors:** Tomoko Nakanishi, Sara Pigazzini, Frauke Degenhardt, Mattia Cordioli, Guillaume Butler-Laporte, Douglas Maya-Miles, Beatriz Nafría-Jiménez, Youssef Bouysran, Mari Niemi, Adriana Palom, David Ellinghaus, Atlas Khan, Manuel Martínez-Bueno, Selina Rolker, Sara Amitano, Luisa Roade Tato, FinnGen, The COVID-19 Host Genetics Initiative, Francesca Fava, Christoph D. Spinner, Daniele Prati, David Bernardo, Federico Garcia, Gilles Darcis, Israel Fernández-Cadenas, Jan Cato Holter, Jesus Banales, Robert Frithiof, Krzysztof Kiryluk, Stefano Duga, Rosanna Asselta, Alexandre C Pereira, Manuel Romero-Gómez, Luis Bujanda, Johannes R. Hov, Isabelle Migeotte, Alessandra Renieri, Anna M. Planas, Kerstin U. Ludwig, Maria Buti, Souad Rahmouni, Marta E. Alarcón-Riquelme, Eva C. Schulte, Andre Franke, Tom H Karlsen, Luca Valenti, Hugo Zeberg, J. Brent Richards, Andrea Ganna

## Abstract

**Background:** There is considerable variability in COVID-19 outcomes amongst younger adults—and some of this variation may be due to genetic predisposition. We characterized the clinical implications of the major genetic risk factor for COVID-19 severity, and its age-dependent effect, using individual-level data in a large international multi-centre consortium.

**Method:** The major common COVID-19 genetic risk factor is a chromosome 3 locus, tagged by the marker rs10490770. We combined individual level data for 13,424 COVID-19 positive patients (N=6,689 hospitalized) from 17 cohorts in nine countries to assess the association of this genetic marker with mortality, COVID-19-related complications and laboratory values. We next examined if the magnitude of these associations varied by age and were independent from known clinical COVID-19 risk factors.

**Findings:** We found that rs10490770 risk allele carriers experienced an increased risk of all-cause mortality (hazard ratio [HR] 1·4, 95% confidence interval [CI] 1·2–1·6) and COVID-19 related mortality (HR 1·5, 95%CI 1·3–1·8). Risk allele carriers had increased odds of several COVID-19 complications: severe respiratory failure (odds ratio [OR] 2·0, 95%CI 1·6-2·6), venous thromboembolism (OR 1·7, 95%CI 1·2-2·4), and hepatic injury (OR 1·6, 95%CI 1·2-2·0). Risk allele carriers ≤ 60 years had higher odds of death or severe respiratory failure (OR 2·6, 95%CI 1·8-3·9) compared to those > 60 years OR 1·5 (95%CI 1·3-1·9, interaction p-value=0·04). Amongst individuals ≤ 60 years who died or experienced severe respiratory COVID-19 outcome, we found that 31·8% (95%CI 27·6-36·2) were risk variant carriers, compared to 13·9% (95%CI 12·6-15·2%) of those not experiencing these outcomes. Prediction of death or severe respiratory failure among those ≤ 60 years improved when including the risk allele (AUC 0·82 vs 0·84, p=0·016) and the prediction ability of rs10490770 risk allele was similar to, or better than, most established clinical risk factors.

**Interpretation:** The major common COVID-19 risk locus on chromosome 3 is associated with increased risks of morbidity and mortality—and these are more pronounced amongst individuals ≤ 60 years. The effect on COVID-19 severity was similar to, or larger than most established risk factors, suggesting potential implications for clinical risk management.

**Funding:** Funding was obtained by each of the participating cohorts individually.

## Introduction

The COVID-19 pandemic has led to the death of millions of individuals and the largest economic contraction since the Great Depression^1^. The clinical outcomes of COVID-19 are remarkably variable, such that some individuals remain asymptomatic^2^, while others develop severe COVID-19 with systemic inflammation, respiratory failure or death. This variability in outcome creates difficulties in clinical management when estimating who is at risk of severe disease and may develop a need for intensive care. Furthermore, recent guidelines suggest risk stratification should be considered when deciding upon prophylactic treatment algorithm and priority for vaccination^3^.

Some of this variation in COVID-19 behavior has been attributed to risk factors such as age^4^, sex^4^, comorbidities^5^, socioeconomic factors^6^ and genetic variants in the SARS-CoV-2 genome.^7^ While the main risk factor for severe outcomes is age, which increases exponentially after age 60^5^, some younger individuals experience severe COVID-19 outcomes and death. The early onset of several common diseases such as breast cancers and myocardial infarction, is disproportionally influenced by human genetic factors^8–10^ and this may also be the case for COVID-19. Several studies have identified and replicated a major genetic risk locus for severe COVID-19^11–13^ in the human genome. This genetic risk locus harbors a cluster of genes on chromosome 3, in which the true causal variant is still unknown. The single nucleotide polymorphism (SNP) rs10490770 serves as a marker for this genetic risk (as well as other SNPs in linkage disequilibrium^14^) and approximately 15% of individuals of European ancestry carry the C risk allele^15^. However, the clinical relevance of this locus, and its potential age-dependent impact, is unknown.

We therefore assembled individual-level COVID-19 clinical and human genomic data in a large international consortium of 17 cohorts in nine countries (Belgium, Brazil, Canada, Germany, Italy, Norway, Spain, Sweden, and UK) to assess the relationship between the chromosome 3 genetic risk with COVID-19 severity, complications and mortality. We next tested the age-dependent effects of this locus on COVID-19 outcomes. Last, in order to assess the relative importance of this locus, we compared its ability to predict COVID-19 outcomes to that of other established clinical risk factors.

## Methods

### Study participants

We gathered clinical and genomic data from 13,424 COVID-19 cases (6,689 of whom were hospitalized) with genetic information available, harmonizing individual-level data from 17 studies. COVID-19 cases were defined as individuals having at least one confirmed SARS-CoV-2 viral nucleic acid amplification test from relevant biologic fluids, or whose SARS-CoV-2 status was confirmed by ICD-10 codes, using codes U071 and/or U072. We combined data from hospital-based studies which recruited participants after COVID-19 outbreak, and a population-based biobank in which recruitment was not dependent upon COVID-19 status. Detailed information for each individual study is described in the online supplement.

### Statistical analysis

In order to tag the chromosome 3 locus, we selected the SNP rs10490770, which was most significantly associated with hospitalization in the COVID-19 genome-wide association study (GWAS) from the COVID-19 Host Genetics Initiative, since this is the largest genome-wide association study meta-analysis of COVID-19 severity^13^ (cases / controls = 12,888 / 1,295,966). Each participating study performed genotyping and imputation separately following a recommended quality control pipeline^16^. Detailed methods describing genotyping and imputation are available in the online supplement. Ancestry was inferred by performing projection onto the principal component analysis (PCA) space from the 1000G^17^ Phase 3 population using HapMap3 SNPs^18^ with minor allele frequency > 1% (detailed methods are in the online supplement) (Supplementary Table 1, Supplementary Figure 1). To test the association between rs10490770 and all phenotypes above, we applied a dominant model by grouping participants into two groups according to their genotype at rs10490770 – C is the allele associated with COVID-19 severity; those with TC genotype or CC genotype were labeled as carriers and those with TT genotype were labeled as non-carriers. We chose this model because it had the lowest Akaike Information Criterion (AIC), compared to additive and recessive models (see the online supplement for detail, Supplementary Table 2), in a logistic regression for death or severe respiratory failure outcome (defined below). All analyses were performed separately for each ancestry group. Because the sample size in non-Europeans was limited, we reported the results from European descent as main analyses, but also reported the results from non-European ancestry individuals are in the supplement. All analyses were based on mixed-effects model adjusted for age, sex and the first five genetic principal components (PCs) as fixed effects and study groups were also included as random effects to account for the study variability. Five study groups, mostly reflecting the country of origin of the study, were created by combining small participating studies with few cases and controls to reduce the risk of collinearity (detail is described in the online supplement). We further estimated the frequency of rs10490770 risk allele carrier status from the population frequencies reported in external database (the Genome Aggregation Database v 3·1 [gnomAD^15^]), assuming this variant follows Hardy-Weinberg equilibrium.

**Table 1.** Patients’ characteristics

|  |  |  |
| --- | --- | --- |
|  | **Hospitalized** | **Total** |

|  | (N=6,689) | (N=13,424) |
| --- | --- | --- |
| **Female** | 2,650 (39.6%) | 6,344 (47.3%) |
| **Age (years)\*** | 64.9 (14.7) | 63.8 (12.7) |
| **Ancestry** |  |  |
| European | 5,601 (83.7%) | 11,658 (86.8%) |
| South Asian | 109 (1.6%) | 388 (2.9%) |
| African | 233 (3.5%) | 421 (3.1%) |
| Others | 185 (2.8%) | 275 (2.0%) |
| East Asian | 59 (0.9%) | 108 (0.8%) |
| Admixed American | 502 (7.5%) | 574 (4.3%) |
| **ICU admission** | 1,622 (25.0%) | 1,622 (12.4%) |
| **Death Status** |  |  |
| Survived | 4,437 (78.4%) | 10,951 (90.0%) |
| Deceased | 1,223 (21.6%) | 1,223 (10.0%) |
| **Respiratory failure** |  |  |
| Severe respiratory failure | 1,644 (32.0%) | 1,644 (14.7%) |
| Oxygen supplementation | 1,641 (32.0%) | 1,641 (14.6%) |
| **Hepatic injury** | 447 (10.0%) | 447 (4.1%) |
| **Cardiovascular complications** | 799 (17.1%) | 804 (7.5%) |
| **Kidney injury** | 1,095 (22.5%) | 1,097 (9.6%) |
| **Venous thromboembolism** | 286 (6.9%) | 287 (2.7%) |

Age\*: Mean (SD), % was calculated amongst those with complete information. The missing rates per each study are listed in Supplementary Table 4.

**Table 2.** Age and risk allele carrier status by COVID-19 severity outcomes

|  | Death or severe respiratory failure | COVID positive but no oxygen supplementation |  |
| --- | --- | --- | --- |
|  |  | Hospitalized only | All |
| **All** |  |  |  |
| carrier | 25.1% [23.2; 27.1] (483) | 16.2% [14.4; 18] (258) | 13.8% [13; 14.6] (974) |
| non-carrier | 74.9% [72.9; 76.8] (1442) | 83.8% [82; 85.6] (1,339) | 86.2% [85.4; 87] (6,081) |
| Total | 100% (1,925) | 100% (1,597) | 100% (7,055) |
| **Age $\leq 60$ years old** |  |  |  |
| carrier | 31.8% [27.6; 36.2] (143) | 14.5% [11.2; 18.5] (51) | 13.9% [12.6; 15.2] (366) |
| non-carrier | 68.2% [63.8; 72.4] (307) | 85.5% [81.5; 88.8] (301) | 86.1% [84.8; 87.4] (2,273) |
| Total | 100% (450) | 100% (352) | 100% (2,639) |
| **Age $> 60$ years old** |  |  |  |
| carrier | 23.1% [21; 25.3] (340) | 16.6% [14.7; 18.8] (207) | 13.8% [12.8; 14.8] (608) |
| non-carrier | 76.9% [74.7; 79] (1,135) | 83.4% [81.2; 85.3] (1,038) | 86.2% [85.2; 87.2] (3,808) |
| Total | 100% (1,475) | 100% (1,245) | 100% (4,416) |

Frequency of rs10490770 risk variant carriers in individuals of European descent stratified by age and COVID-19 severe outcomes. [95%CI] (Sample size)

**Figure 1:**
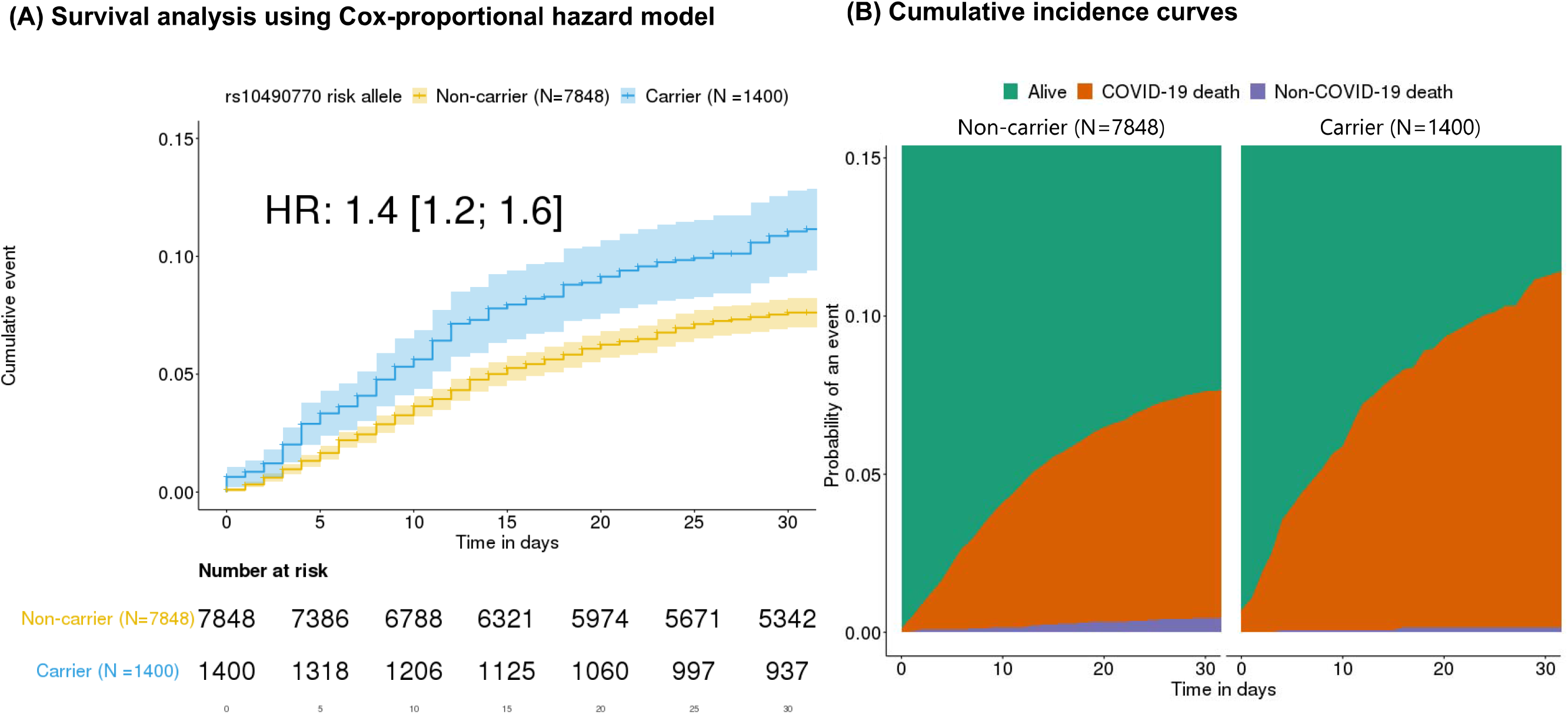
Associations with mortality. The results described here were restricted to 9,248 COVID-19 patients of European ancestry with available follow-up and cause of death information. (A) Kaplan-Meier curves stratified by rs10490770 risk allele carrier status. (Carriers: N=1,400 vs non-carriers: N=7,848). Hazard ratios (HR) were calculated by adjusting for age, sex, genetic PCs 1 to 5 as fixed effects, and groups indicating participating studies as random effects. (B) Cumulative incidence curves for COVID-19 related death and COVID-19 unrelated death amongst the same individuals as (A).

### Association with mortality

The hazard ratio (HR) for all-cause mortality was estimated by Cox proportional hazard models using the “coxme v2·2-16” R package. Individuals entered the follow-up when diagnosed with COVID-19 or if a diagnosis date was missing, the date when they were hospitalized or when their symptoms started. They were considered as an event at the date of death and censored at the last date of follow-up (details are described in the online supplement). We additionally performed competing risk analyses to estimate the sub-distribution hazard ratio for COVID-19 related mortality using the “cmprsk v2·2-10” R package, which accounts for the competing risk of non-COVID-19 related death: i.e. individuals who did not die of COVID-19 but died due to other causes (e.g. cancer). In the competing risk model, study groups were considered as fixed effects. Survival analyses were restricted to study participants with available follow-up and cause of death information (N=9,248). Cause of death was defined by doctor-diagnoses, medical chart reviews or ICD-10 codes (details are described in the online supplement).

### Association with COVID-19 severity and complications

To understand the clinical implications of the chromosome 3 locus, we fit mixed-effects regression models to assess the association of rs10490770 risk allele [C] carrier status with three types of COVID-19 outcomes: COVID-19 severity, COVID-19 complications and laboratory values. To do so, we defined three COVID-19 severity outcomes, with appropriate control definitions amongst SARS-CoV-2 positive individuals. 1) hospitalization; 2) intensive care unit (ICU) admission and 3) death or severe respiratory failure. Hospitalization cases were COVID-19 cases admitted to the hospital, whereas controls were individuals who did not experience hospitalization. ICU cases were those COVID-19 cases admitted to the ICU and controls were individuals who did not experience hospitalization. To assess potential selection bias, we also repeated the analyses using only individuals who were hospitalized. In these analyses, controls were defined as those who were hospitalized, but not admitted to the ICU. Death or severe respiratory failure cases were defined as individuals who died or required respiratory support (intubation, continuous positive airway pressure, Bilevel Positive Airway Pressure, or continuous external negative pressure, Optiflow/high flow Positive End Expiratory Pressure Oxygen), had ICD-10 codes for acute respiratory distress syndrome (ARDS) or acute respiratory failure (“J80”, “J9600”,”J9609”,”Z991”), or OPCS codes of the use of ventilator (“E851”,”E852”). Controls for the death or severe respiratory failure cases were defined as those requiring no oxygen therapy and who were alive.

We next defined five COVID-19 related complications, which were diagnosed at hospital. These included: 1) Severe respiratory failure, which was defined by the use of respiratory support or individuals with administrative codes for ARDS, respiratory failure or ventilatory support as described above; 2) Hepatic injury was defined as individuals with at least one of the following: doctor-diagnosed hepatic complications, highest alanine aminotransferase > 3 times upper limit of normal (ULN), or ICD-10 codes for acute hepatic failure (“K720”); 3) Cardiovascular complications were defined by at least one of the following: doctor-diagnosed acute myocardial infarction (AMI) or stroke, highest troponin T or troponin I > ULN, or ICD-10 codes for AMI or stroke (“I21*”,”I61”, “I62”, “I63”, “I64”, “I65”, “I66*”).; 4) Kidney injury was defined by at least one of the following: doctor-diagnosed acute kidney injury (AKI), highest creatinine > 1·5 times ULN, or ICD-10 codes for AKI (“N17*”); 5) Venous thromboembolism (VTE) was defined by at least one of the following: doctor-diagnosed pulmonary embolism (PE) or deep venous thrombosis (DVT), or ICD-10 codes for PE or DVT (“I26*”, “I81”, “I82*”). Controls for severe respiratory failure were defined as those requiring no oxygen therapy and who were alive, whereas controls for other complications were defined as those who did not meet the corresponding case criteria and were alive.

Last we considered the laboratory values of complete blood count and biochemistry tests available at hospital (Supplementary Table 3). To test the association with the chromosome 3 locus we used the highest or lowest value recorded per individual^19–23^. We selected the lowest value for lymphocyte counts and otherwise highest value. This is because we were interested in using these laboratory values as a proxy of COVID-19 severity. Definitions and quality control of laboratory values and specific codes are described in the online supplement (Supplementary Figure 2).

**Table 3:** Risk prediction performance for death or severe respiratory failure

| Age range | Model | AUC † | AUC p-value* | NRI † | NRI p-value* |
| --- | --- | --- | --- | --- | --- |
| All<br>Cases = 834<br>Controls = 6,454 | Baseline | 0.77 [0.75; 0.78] | - | - | - |
| | Baseline and rs10490770 | 0.78 [0.76; 0.79] | $1.8 \times 10^{-4}$ | 0.19 [0.13; 0.25] | $2.0 \times 10^{-10}$ |
| Ages $\leq 60$<br>Cases = 128<br>Controls = 2,348 | Baseline | 0.82 [0.78; 0.86] | - | - | - |
| | Baseline and rs10490770 | 0.84 [0.81; 0.88] | $1.6 \times 10^{-2}$ | 0.45 [0.29; 0.62] | $6.5 \times 10^{-8}$ |

Only individuals with complete information of clinical risk factors and genotype were included. Baseline model includes age, sex, BMI, smoking status (ever-smoker vs never-smoker), cancer, chronic kidney disease, chronic obstructive pulmonary disease (COPD), chronic heart failure, transplantation, and diabetes mellitus. \*p-values were calculated by comparing baseline model and baseline and rs10490770 model. †: [95%CI]

**Figure 2:**
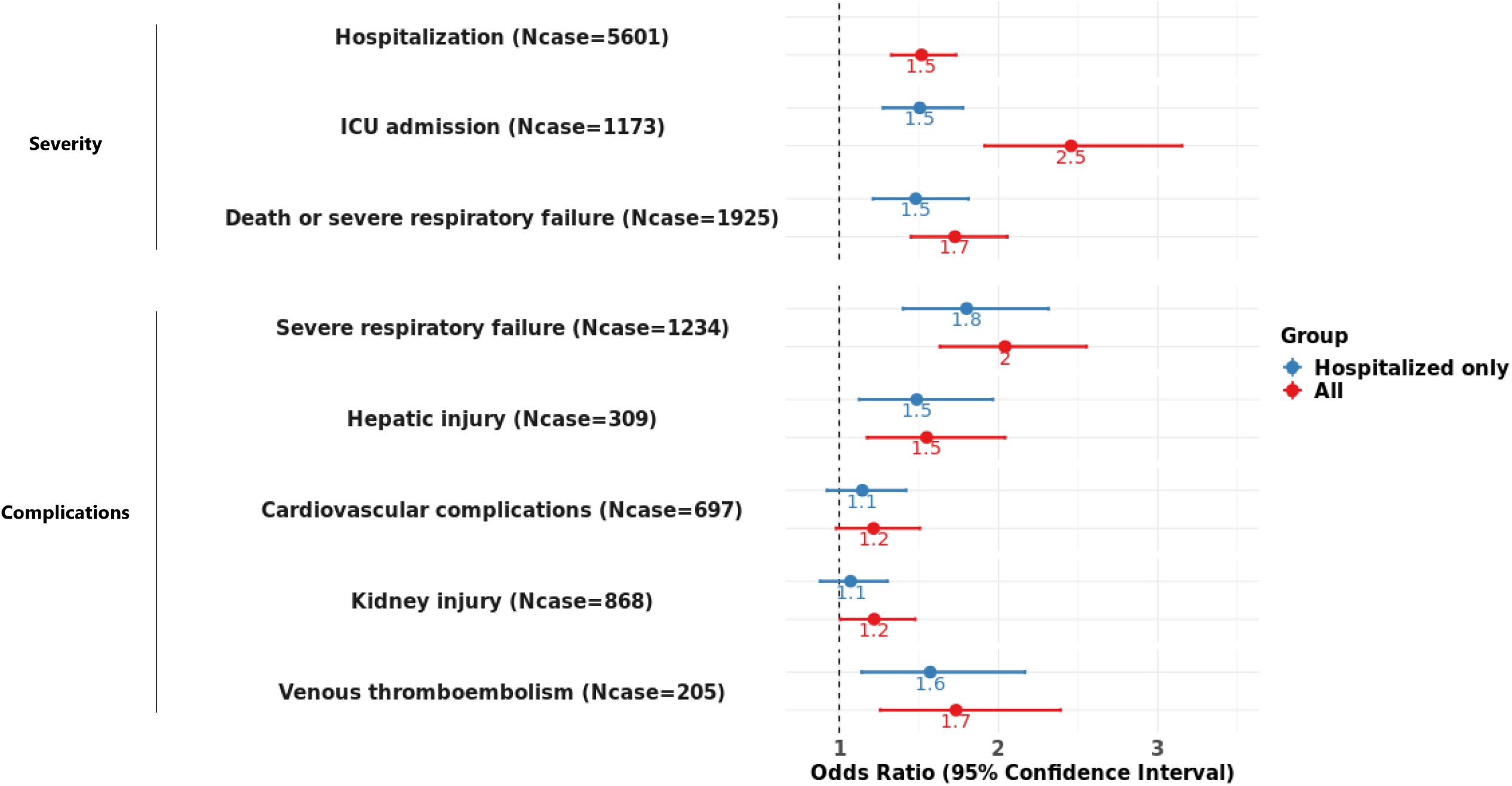
Associations between rs10490770 risk allele carrier status and COVID-19 severity and complications. The results described here were restricted to COVID-19 patients of European ancestry. Logistic regressions were fit to assess the associations of rs10490770 risk allele carrier status with COVID-19 severity and complications, adjusting for age, sex, genetic PCs 1 to 5 as fixed effects, and groups indicating participating studies as random effects. Red: All participants (N=11,658) Blue: Hospitalized participants only (N=5,601) The case counts demonstrated here are from the data in all individuals.

### Age-dependent associations with COVID-19 severity

We evaluated the age-dependent effects of the risk allele carrier status on COVID-19 three severity phenotypes by performing two sets of analyses: 1) linear regressions between age at diagnosis and risk allele carrier status amongst severe cases, adjusting for the same covariates as the main analyses, and 2) adding a carrier status by age interaction term in the main regression models. Age was not dichotomized in these analyses. We also stratified participants by age ≤60 or >60 years and repeated the same logistic regressions, as well as we estimated the frequency of the risk allele carriers in the two age groups. We used 60 years as a cut-point for age-stratified analyses, because COVID-19 case fatality rates increased markedly after this age^24,25^.

### Associations with COVID-19 severity stratified by established clinical risk factors

In order to compare the association of rs10490770 risk allele carrier status with other risk factors, we similarly stratified participants by BMI ≥30 kg/m^2^ (a definition of obesity^26^), smoking (ever-smoker vs never-smoker), cancer, chronic kidney disease, chronic obstructive pulmonary disease (COPD), chronic heart failure, transplantation, and diabetes mellitus (DM), all of which were curated as established clinical risk factors for severe illness of COVID-19 according to the Centre for Disease Control website^26^. All of the eight risk factors were defined by doctor-diagnoses, medical chart reviews or ICD-10 codes (details are described in the online supplement). We then tested the difference of the magnitude of the associations of the risk allele carrier status compared to the eight clinical risk factors. Clinical risk factors stratified analysis and prediction assessment (described below) were restricted to individuals with complete information for demographics, clinical risk factors and rs10490770 genotype information (N=7,919). The majority of this subset were from UK Biobank (N=7,461), and only 50 individuals were included from the first discovery GWAS^11^.

### Risk prediction compared to established clinical risk factors

To better understand the prediction improvement by adding of the chromosome 3 genetic risk in addition to the eight clinical risk factors, we performed multivariate regressions in individuals with complete information as described above (N=7,919). We evaluated whether the rs10490770 risk allele improved the risk prediction discrimination for severe COVID-19 outcomes by calculating the area under receiver operation curve (AUC) and the continuous net reclassification improvement (NRI) using “pROC v1·16·2” and “PredictABEL v1·2-4” R packages.

### Meta-analyses

As secondary analyses, we meta-analyzed the results with non-European ancestries and two external cohorts for which we did not have access to individual-level data; FinnGen and Columbia University COVID-19 Biobank (CUB). This resulted in a total study population of 14,620 individuals with COVID-19. An inverse-variance weighted meta-analyses were performed under a fixed effect and random effects models using the “meta v4·16-1” R package when the appropriate phenotypes were available and case counts, control counts, and the rs10490770 risk allele carrier counts were larger than ten in each cohort.

### Sensitivity analysis

Adjusting for participating studies may lead to reduced statistical power, given that some studies had only severe cases or had disproportional case-control ratio. To alleviate the collinearity issue, we grouped some small studies to account for study variability. This may not fully account for between study variability. Thus we performed two sets of sensitivity analyses where we included, 1) only five genetic PCs without including the study of origin as random or fixed effects, and 2) including all participating studies either as fixed or random effects. Next, we performed the same analyses using UK Biobank (UKB) to provide estimates which are more representative of general population, since this is not a COVID-19 specific cohort. We also tried binning by different cut-offs for age-stratified analyses. In order to understand if results could have been influenced by related individuals within the samples, we selected one individual from a pair of relatives with PI-HAT (proportion of identity by descent calculated by PLINK^27^) >0·1875 (meaning between second and third-degree relatives) and repeated the main analyses.

### Role of the funding source

The funding sources had no role in study design; in the collection, analysis, and interpretation of data; in the writing of the report; and in the decision to submit the paper for publication.

## Results

### Study participants

We collected and harmonized individual-level data from 13,424 COVID-19 patients diagnosed with COVID-19 from February 5^th^, 2020 to January 2^nd^, 2021. Table 1 illustrates the participants’ demographic and clinical characteristics. The majority of participants were of European descent (11,658; 86·8%). However, important numbers of non-European descent individuals were also included in meta-analyses: 388 (2·9%) were South Asian ancestry and 574 (4·3%) were Admixed-American ancestry. 6,689 were hospitalized, amongst whom 1,622 (25·0%) were admitted to the ICU. 1,223 (21·6%) died following COVID-19 diagnosis and 1,644 (31·9%) met the criteria for severe respiratory failure. Clinical information was obtained with different degrees of completeness across studies. A detailed description of study-specific demographic, clinical characteristics and their missingness rates is provided in Supplementary Table 4.

### Risk allele frequency

According to the population frequencies in gnomAD^15^, we estimate that 15·6% of individuals of European descent carry at least one rs10490770 C allele, as well as 10·0% of Latinx/Admixed-American, 2·4% of African/African-American, 62·0% of South Asians and 0·4% of East Asians. In our study the carrier frequency was 16·2% amongst individuals of European descent in our cohort.

### Association with mortality

Risk allele carriers at rs10490770 had a higher HR for all-cause mortality compared to non-carriers (HR 1·4, 95%CI 1·2–1·6, p=1·1×10^−4^, dead / alive = 832 / 8,416) over a median follow-up duration of 45 days (interquartile range [IQR] 21-70 days) (Figure 1A). A competing risk model to estimate the HR for COVID-19-related death while accounting for non-COVID-19-related deaths estimated a similar HR for COVID-19 related mortality (HR 1·5, 95%CI 1·3-1·8, p=2·7×10^−6^, dead / alive = 720 / 8,416) (Figure 1B). The association with mortality was reduced when the analysis was restricted to hospitalized individuals (HR for all-cause mortality 1·2, 95%CI 1·0–1·4, p=0·061, dead / alive = 832 / 2,796, and HR for COVID-19 related mortality 1·3, 95%CI 1·1-1·5, p=4·2×10^−3^, dead / alive = 720 / 2,796).

### Associations with COVID-19 severity

We confirmed that risk allele carrier status at rs10490770 was significantly associated with hospitalization (OR 1·5, 95%CI 1·3-1·7, p=9·1×10^−10^, cases / controls = 5,601 / 5,997). A stronger effect was observed for ICU admission (OR 2·5, 95%CI 1·9-3·2, p=1·9×10^−12^, cases / controls = 1,173 / 6,004) and death or severe respiratory failure (OR 1·7, 95%CI 1·5-2·1, p=7·7×10^−10^, cases / controls = 1,925 / 7,055) (Figure 2, Supplementary Table 5). Restricting analyses to hospitalized individuals, we observed consistent results, some of which were with diminished effect sizes (Figure 2, Supplementary Table 5). For instance, a significant reduction in effect size was observed in OR for ICU admission (OR 1·5, 95%CI 1·3-1·8, p=1·4×10^−6^, cases / controls = 1,173 / 4,428)

We next explored the association of the rs10490770 risk allele with laboratory values, which are known to be associated with the severity of COVID-19^19–23^. rs10490770 risk allele carrier status was associated with the worst value for each of these laboratory values at hospital (e.g. lactate dehydrogenase: 0·24 SD increase, p=2·8×10^−6^, D-dimer: 0·15 SD increase, p=3·6×10^−3^ and interleukin-6: 0·18 SD increase, p=6·3×10^−3^; Supplementary Figure 3, Supplementary Table 3).

**Figure 3:**
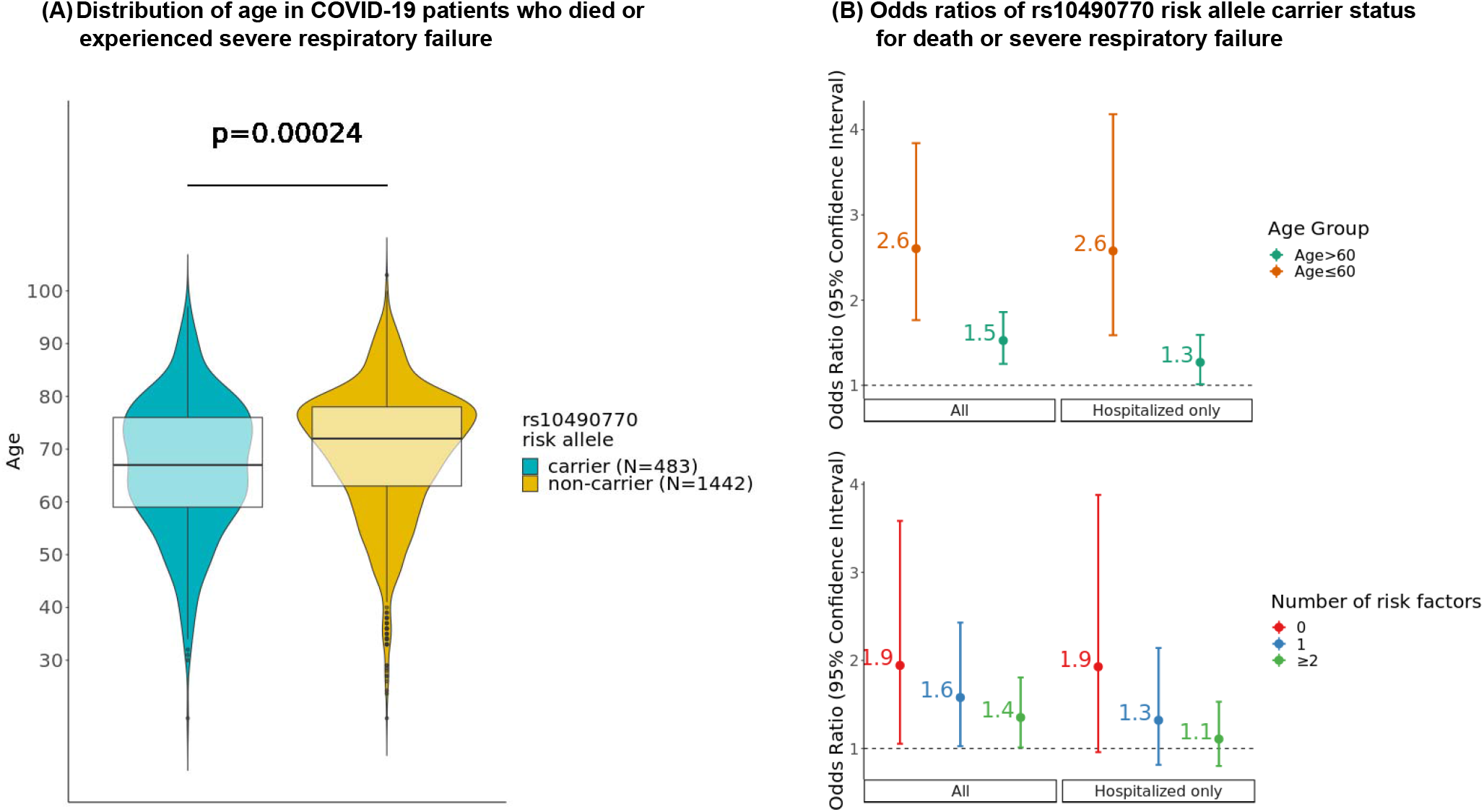
Influence of age and clinical risk factors for the effect of rs10490770 risk allele carrier status on death or severe respiratory failure. (A) Age distribution in COVID-19 patients of European ancestry who died or experienced severe respiratory failure (N=1,925). Median (IQR) age was 67 (63-78) years in carriers (N=438) and 72 (59-76) years in non-carriers (N=1,442). (B) Logistic regressions between rs10490770 risk allele carrier status and death or severe respiratory failure. Regressions were performed within subgroups stratified by age (age ≤ 60 years and age > 60 years) (Cases / Controls = 1,925 / 7,055) or by the number of established risk factors (0, 1, or ≥2); BMI≥30, smoking, cancer, chronic kidney disease, chronic obstructive pulmonary disease (COPD), chronic heart failure, transplantation, and diabetes mellitus (Cases / Controls = 834 / 6,454).

### Associations with COVID-19 complications

Risk allele carrier status at rs10490770 was associated with multiple COVID-19-related severe complications (Figure 2). These included severe respiratory failure (OR 2·0, 95%CI 1·6-2·6, p=4·3×10^−10^, Cases / Controls = 1,234 / 7,055), VTE (OR 1·7, 95%CI 1·3-2·4, p=5·7×10^−3^, Cases / Controls = 205 / 8,914) and hepatic injury (OR 1·5, 95%CI 1·2-2·0, p=1·9×10^−3^, Cases / Controls = 309 / 9,190). No significant effect was observed for cardiovascular complications (OR 1·2, 95%CI 1·0-1·5, p=0·075, Cases / Controls = 697 / 8,611), although this might be due to lack of statistical power to detect such effects. Similar results were observed when restricting to hospitalized patients (Figure 2; Supplementary Table 5), indicating that the effect of rs10490770 on severe COVID-19 complications was not simply explained by the higher hospitalization rate among the carriers.

### Age-dependent associations with COVID-19 severity

We explored the age-dependent effects of rs10490770 risk allele carrier status on severe COVID-19 outcomes in individuals of European descent. Amongst severe patients who died or had severe respiratory failure, rs10490770 risk allele carriers were on average 2·3 (95%CI 1·1-3·5) years younger than non-carriers (p=2·4×10^−4^, N=1,925, Figure 3A; Supplementary Table 5). Stratifying by age, we found that amongst those who were ≤ 60 years, risk allele carrier status had markedly increased odds of death or severe respiratory failure (OR 2·6 95%CI 1·8-3·8), whereas risk allele carrier status had more modest effects amongst those >60 years with an OR of 1·5 (95%CI 1·3-1·9, p-value interaction=0·043, Figure 3B, Supplementary Table 5-6). Amongst all participants ≤ 60 years who died or experienced a severe respiratory COVID-19 outcome, we found that 31·8% (95%CI 27·6-36·2%) were rs10490770 risk variant carriers, compared to 13·9% (95%CI 12·6-15·2%) of those who did not experience severe disease (Table 2). When considering other severity phenotypes, such as hospitalization and

ICU admission, we observed that risk allele carriers tend to be younger than non-carriers. However, we did not detect a different effect in the association between rs10490770 risk allele carriers and these additional severity phenotypes amongst those who were ≤60 vs >60 years old. This could be attributed to the heterogeneity of the criteria of hospitalization or ICU admission, or case-control imbalance in some participating studies.

### Associations with COVID-19 severity stratified by established clinical risk factors

We studied how the effects of rs10490770 risk allele carrier status on COVID-19 severity varied by other established clinical risk factors. Amongst individuals with no risk factors (BMI ≥ 30, smoking, cancer, chronic kidney disease, chronic obstructive pulmonary disease, heart failure, transplantation, and DM) prior to COVID-19, risk allele carriers had an OR of 1·9 for death or severe respiratory failure (95%CI 1·1-3·6), whereas risk allele carrier status had more modest effects amongst those with one medical condition (OR 1·6, 95%CI 1·0-2·4) and more than one medical conditions (OR 1·4, 95%CI 1·0-1·8) (p-value for interaction=0·087; Figure 3B, Supplementary Table 7).

### Risk prediction compared to established clinical risk factors

We compared the risk discrimination conferred by the rs10490770 risk allele on COVID-19 severity with that observed for other established COVID-19 risk factors. To do so, we used multivariate regression in individuals of European ancestry with complete ascertainment of clinical risk factors. rs10490770 risk allele carrier status was independent of other risk factors (Figure 4A, Supplementary Table 8) when examining the association with death or severe respiratory failure (OR 2·0, 95%CI 1·7-2·4, p=4·7×10^−13^, frequency of risk allele carriers 14·6%, Cases / Controls = 834 / 6,454). The effect sizes were comparable, or larger, than those of other known risk factors such as DM (OR 1·9, 95%CI 1·6-2·4, p=6·2×10^−12^, frequency of DM 12·3%). Stronger effects were observed amongst individuals ≤60 years (risk allele carrier status: OR 4·0, 95%CI 2·6-6·2, p=1·3×10^−10^, Cases / Controls = 128 / 2,348) relative to DM (OR 2·3, 95%CI 1·3-4·2, p=5·7×10^−3^, frequency of DM: 5·5%) (Figure 4A, Supplementary Table 8).

**Figure 4:**
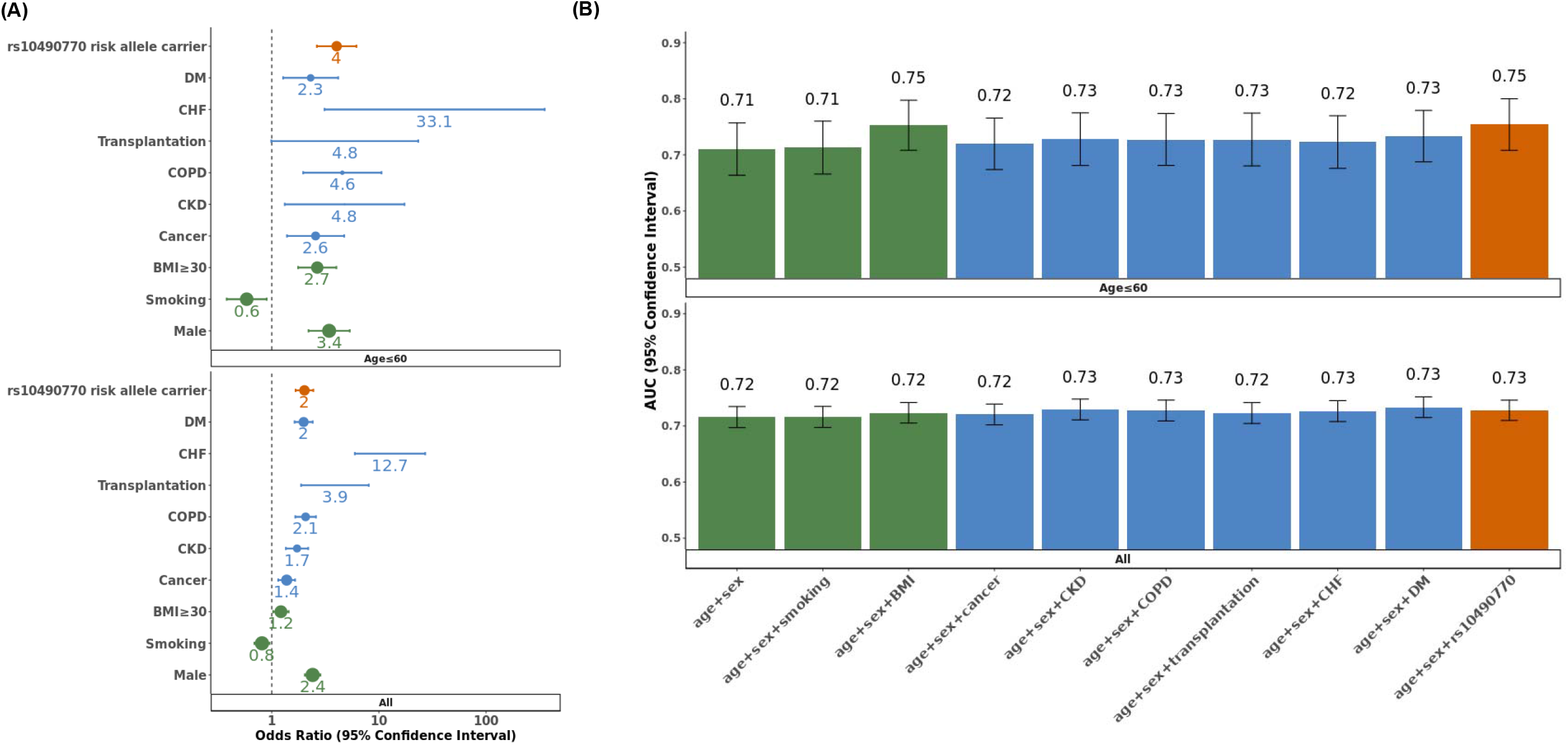
Multivariate regression models and risk prediction estimates of COVID19 death or severe respiratory failure. Multivariate regression analyses for death or severe respiratory failure were restricted to European-ancestry individuals with complete information of demographic variables (green), comorbidities (blue) and rs10490770 risk allele status (red). (N=7,288 for all and N = 2,476 for Age ≤ 60), CKD: chronic kidney disease, COPD: chronic obstructive pulmonary disease, CHF: chronic heart failure, DM: diabetes mellitus. (A) Forest plots comparing odds ratios from multivariate regression models. The size of each dot represents the frequency of the risk factors. (B) Comparison of AUCs of predictions for COVID-19 outcomes. rs10490770 risk allele and non-genetic clinical risk factors were included separately in addition to age and sex in multivariate regression models

Consistent with the results from multivariate regression, adding rs10490770 genotype to non-genetic risk factors improved discrimination for death or severe respiratory failure amongst ≤ 60 years (AUC: 0·82 vs 0·84, p=0·016 and NRI 0·45, p=6·5×10^−8^, Table 3), and the performance of risk discrimination was similar to, or better than, most of established risk factors included in the study (Figure 4B, Supplementary Table 9).

### Meta-analyses

We meta-analyzed the European ancestry results presented above with those of non-European ancestry participants and two external cohorts. We confirmed similar effects in the associations with mortality (Supplementary Figure 4), COVID-19 severity (Supplementary Figure 5), COVID-19 complications (Supplementary Figure 6) and age-dependent effects (Supplementary Figure 7). Given the small sample size of non-European participants, we lacked sufficient statistical power to investigate whether the association between rs10490770 risk allele carriers and COVID-19 outcomes was different when comparing individuals of non-European and European ancestry.

### Sensitivity analysis

Last, we performed several sensitivities analyses to evaluate the robustness of our results. First, we removed the study variables from the covariates and instead included the top five PCs (Supplementary Table 10-11). Second, we included participating studies themselves either as fixed or random effects (Supplementary Table 10-11). Third, we restricted to individuals of European descent from UKB, a cohort which was not developed to study COVID-19 and thus is less prone to selection bias. These UKB analyses generated similar results (Supplementary Table 12). Fourth, we explored different cut-offs for age-stratified analyses (Supplementary Table 13). Last, we excluded related individuals (Supplementary Table 14). All sensitivity analyses were consistent with the results from the main analyses.

## Discussion

Combining individual-level data from 13,424 individuals ascertained for COVID-19 outcomes from 17 cohorts in nine countries, we found that the major genetic risk factor for severe COVID-19 on chromosome 3 was strongly associated with COVID-19 related mortality and clinical complications such as respiratory failure and venous thromboembolism. Effect sizes were considerably larger in individuals ≤60 years and this genetic risk factor was similar in magnitude, and often more common, than most established clinical risk factors. These findings suggest that this genetic variant should be considered in risk stratification for COVID-19 outcomes.

The risk allele is common. We estimated that 15·6% of individuals of European ancestry are risk allele carriers at rs10490770. Further, 10·0% of Latinx/Admixed Americans, 2·4% of African/African-American, 62·0% of South Asians and 0·4% of East Asians are risk allele carriers^15^. Consequently, a large proportion of individuals carry this risk factor.

The effect of carrying the risk allele on COVID-19 severity was stronger in younger individuals. First, amongst those ≤60 years, the odds of death or severe respiratory failure increased 2·6-fold for risk allele carriers. We found that 32% of individuals ≤60 years who died, or experienced severe respiratory failure, were risk allele carriers, compared to 14% of individuals not requiring supplemental oxygen. Second amongst individuals who died, or experienced severe respiratory failure, risk allele carriers were on average 2·3 years younger than non-carriers. Last, the risk discrimination for death and severe respiratory COVID-19 provided by the risk allele was similar to, or larger than, established clinical risk factors in individuals ≤60 years. Other common diseases have also demonstrated larger effects of genetic risk factors at younger age^8,9^. Genetic risk factors are often clinically valuable for risk stratification in younger age groups because the frequency of other established risk factors for COVID-19 are often reduced, while the frequency of the genetic variant remains high. Moreover, this specific variant is not associated with any known COVID-19 risk factor and therefore provide orthogonal information compared to existing risk assessment tools.

Our findings suggest potential implications for clinical risk assessments in three situations. Currently, risk factors such as DM are clinically used in triage to decide if COVID-19 patients require further follow-up. Amongst individuals less than 60 years old, this genetic risk factor has considerably larger effect size and is more common than DM. This suggests that genotyping could help to identify individuals who are at risk for COVID-19 severe outcomes and death, allowing for more tailored treatment and clinical observation. Second, amongst very ill individuals less than 60 years, the genetic risk factor is quite common and may help to explain to patients and families why this individual has become severely ill, while others with the same clinical risk factor profile remain healthy. Last, since SARS-CoV-2 will become endemic in the human population, future public health strategies, including vaccines against novel variants of

SARS-CoV-2, could be targeted to individuals at higher risk of severe outcomes. The major common genetic risk factor for severe COVID-19 could help to ensure individuals at highest risk are prioritized for vaccine programs, thus reducing the overall burden of the disease. The biology of how this chromosome 3 genetic risk has an effect on COVID-19 severity is still unknown. This locus on chromosome 3p21 includes the putative SARS-COV-2 coreceptors; *SCL6A20*^28,29^, *LZTFL1, FYCO1*^30^, and the chemokine receptors; *CCR9*^29^, *CXCR6*^31^, *XCR1*. There are other flanking genes; *CCR1, CCR2 and CCR3*^32–34^, whose involvement in SARS-CoV-2 infection had been suggested and could explain the biology of the striking effect of this genetic risk. Many studies^12,29^ had been trying to pinpoint a or a set of causal genes but the consensus had not been built to date.

This study has important limitations. Each cohort has its own selection bias and ascertainment bias. Several studies were enriched for severe patients, whereas UKB is a non-COVID-19 cohort, with evidence of healthy volunteer bias^35^. Nevertheless, it may be less prone to selection bias than the COVID-19 cohorts. Selection bias is inherent to most COVID-19 observational studies^36^ and this influences the generalizability of the results outside the study populations. To mitigate against these potential issues, we combined data from observational studies with different ascertainment strategies, including national healthcare systems, studies that were established prior to the COVID-19 pandemic and thus recruitment was not dependent upon COVID-19 status and hospital-based studies. This allowed for an increased representation of individuals with severe COVID-19 outcomes. We also provide analyses restricted to hospitalized patients, which is an ascertained, but clinically-relevant population. While we included information from participants who were of non-European ancestry, on-going efforts should enable larger sample sizes in these ancestries to better define the importance of the chromosome 3 risk locus in these contexts. This further emphasizes the importance of developing genomics-enabled studies in individuals of non-European ancestry.

In summary, the major genetic COVID-19 risk locus is common and has large effects on COVID-19 outcomes including mortality. These effects are age-dependent, such that the magnitude of risk increases in younger individuals. These findings suggest potential implications of genetic information in clinical risk management.

## Supporting information

Contributors list

## Data Availability

The harmonized individual-level data of some participating cohorts from Belgium (BeLCovid_2), Brazil (BRACOVID), Italy (COVID19-Host(a)ge_4, GEN-COVID), Spain (COVID19-Host(a)ge_1,2,3, INMUNGEN-CoV2, SPGRX), and Sweden (SweCovid) is under preparation to be deposited at the European Genome-phenome Archive (EGA).
Regarding the data from genetic modifiers for COVID-19 related illness (BelCovid_1), individual level data were acquired and shared with FIMM during the sanitary crisis under an emergency consent and an ethical approval which were specific to this particular project and do not cover deposition to public repositories. Upon contact with Francoise Wilkin, Isabelle Migeotte, or Guillaume Smits, an institutional data transfer agreement can be established and data shared if the aims of data use are covered by ethical approval and patient consent. The procedure will involve an update to the ethical approval, as well as review by legal departments at both institutions and the process will typically take 2-4 months from initial contact.
Regarding the BoSCO study, individual-level genotype and clinical data for purpose of this study were shared with FIMM under a legal, bilateral agreement and were specific to this particular project. Current participant consents and privacy regulations prohibit deposition of individual level data to public repositories. Upon contact with Kerstin Ludwig or Markus M. Nothen, an institutional data transfer agreement can be established and data shared if the aims of data use is covered by ethical approvals and patient consent. The procedure will involve review by legal departments at both institutions and the process will typically take about 2 months from initial contact.
The BQC19 is an Open Science biobank. Instructions on how to access data for individuals from the BQC19 at the Jewish General Hospital site are available here: https://www.mcgill.ca/genepi/mcg-covid-19-biobank. Instructions on how to access data from other sites of the BQC19 are available here: https://www.bqc19.ca/en/access-data-samples.
For the COMRI cohort, data protection legislation does not allow for deposition of individual level data in public repositories. Upon direct contact with Prof Ulrike Protzer (, genetic data) and Dr Christoph Spinner, an institutional data transfer agreement can be established and data will be shared if the aims of data use are covered by ethical approvals and patient consent. The procedure will involve an update to the ethical approval as well as review by legal departments at both institutions and the process will typically take 2-3 months from initial contact.
Regarding the Fondazione IRCCS Milan data (FOGS study), institutional data privacy regulations prohibit deposition of individual level data to public repositories without a specific consent. Participant written consent also does not cover public sharing of data for use for unknown purposes. Upon contact with professor Luca Valenti an institutional data transfer agreement can be established and data shared if the aims of data use are covered by ethical approvals and patient consent. The procedure will involve the request for an amendment to the ethical approvals, as well as review by legal departments at both institutions and the process will typically take 1-2 months from initial contact.
Regarding Norwegian data (NorCoV2), institutional data privacy regulations prohibit deposition of individual level data to public repositories. Participant written consent also does not cover public sharing of data for use for unknown purposes. Upon contact with professor Tom H Karlsen or professor Johannes R. Hov an institutional data transfer agreement can be established and data shared if the aims of data use is covered by ethical approvals and patient consent. The procedure will involve an update to the ethical approvals, as well as review by legal departments at both institutions and the process will typically take 1-2 months from initial contact.
The genetic and phenotype datasets from UK Biobank are available via the UK Biobank data access process (see http://www.ukbiobank.ac.uk/register-apply/).

## Acknowledgement

We thank the patients who volunteered to contribute to all of the participating studies in such difficult times, and the research staffs in every cohort who recruited patients at personal risk.

## Funding

AG has received support by NordForsk Nordic Trial Alliance (NTA) grant, by Academy of Finland Fellow grant N. 323116 and the Academy of Finland for PREDICT consortium N. 340541.

The Richards research group is supported by the Canadian Institutes of Health Research (CIHR) (365825 and 409511), the Lady Davis Institute of the Jewish General Hospital, the Canadian Foundation for Innovation (CFI), the NIH Foundation, Cancer Research UK, Genome Québec, the Public Health Agency of Canada, the McGill Interdisciplinary Initiative in Infection and Immunity and the Fonds de Recherche Québec Santé (FRQS). TN is supported by a research fellowship of the Japan Society for the Promotion of Science for Young Scientists. GBL is supported by a CIHR scholarship and a joint FRQS and Québec Ministry of Health and Social Services scholarship. JBR is supported by an FRQS Clinical Research Scholarship. Support from Calcul Québec and Compute Canada is acknowledged. TwinsUK is funded by the Welcome Trust, the Medical Research Council, the European Union, the National Institute for Health Research-funded BioResource and the Clinical Research Facility and Biomedical Research Centre based at Guy’s and St. Thomas’ NHS Foundation Trust in partnership with King’s College London. The Biobanque Québec COVID19 is funded by FRQS, Genome Québec and the Public Health Agency of Canada, the McGill Interdisciplinary Initiative in Infection and Immunity and the Fonds de Recherche Québec Santé. These funding agencies had no role in the design, implementation or interpretation of this study.

The COVID19-Host(a)ge study received infrastructure support from the DFG Cluster of Excellence 2167 “Precision Medicine in Chronic Inflammation (PMI)” (DFG Grant: “EXC2167”). The COVID19-Host(a)ge study was supported by the German Federal Ministry of Education and Research (BMBF) within the framework of the Computational Life Sciences funding concept (CompLS grant 031L0165). Genotyping in COVID19-Host(a)ge was supported by a philantropic donation from Stein Erik Hagen.

The COVID GWAs, Premed COVID-19 study (COVID19-Host(a)ge_3) was supported by “Grupo de Trabajo en Medicina Personalizada contra el COVID-19 de Andalucia”and also by the Instituto de Salud Carlos III (CIBERehd and CIBERER). Funding comes from COVID-19-GWAS, COVID-PREMED initiatives. Both of them are supported by “Consejeria de Salud y Familias” of the Andalusian Government. DMM is currently funded by the the Andalussian government (Proyectos Estratégicos-Fondos Feder PE-0451-2018).

The Columbia University Biobank was supported by Columbia University and the National Center for Advancing Translational Sciences, NIH, through Grant Number UL1TR001873. The content is solely the responsibility of the authors and does not necessarily represent the official views of the NIH or Columbia University.

The SPGRX study was supported by the Consejería de Economía, Conocimiento, Empresas y Universidad #CV20-10150.

The GEN-COVID study was funded by: the MIUR grant “Dipartimenti di Eccellenza 2018-2020” to the Department of Medical Biotechnologies University of Siena, Italy; the “Intesa San Paolo 2020 charity fund” dedicated to the project NB/2020/0119; and philanthropic donations to the Department of Medical Biotechnologies, University of Siena for the COVID-19 host genetics research project (D.L n.18 of March 17, 2020). Part of this research project is also funded by Tuscany Region “Bando Ricerca COVID-19 Toscana” grant to the Azienda Ospedaliero Universitaria Senese (CUP I49C20000280002). Authors are grateful to: the CINECA consortium for providing computational resources; the Network for Italian Genomes (NIG) (http://www.nig.cineca.it) for its support; the COVID-19 Host Genetics Initiative (https://www.covid19hg.org/); the Genetic Biobank of Siena, member of BBMRI-IT, Telethon Network of Genetic Biobanks (project no. GTB18001), EuroBioBank, and RD-Connect, for managing specimens.

Genetics against coronavirus (GENIUS), Humanitas University (COVID19-Host(a)ge_4) was supported by Ricerca Corrente (Italian Ministry of Health), intramural funding (Fondazione Humanitas per la Ricerca). The generous contribution of Banca Intesa San Paolo and of the Dolce&Gabbana Fashion Firm is gratefully acknowledged.

Data acquisition and sample processing was supported by COVID-19 Biobank, Fondazione IRCCS Cà Granda Milano; LV group was supported by MyFirst Grant AIRC n.16888, Ricerca Finalizzata Ministero della Salute RF-2016-02364358, Ricerca corrente Fondazione IRCCS Ca’ Granda Ospedale Maggiore Policlinico, the European Union (EU) Programme Horizon 2020 (under grant agreement No. 777377) for the project LITMUS-“Liver Investigation: Testing Marker Utility in Steatohepatitis”, Programme “Photonics” under grant agreement “101016726” for the project “REVEAL: Neuronal microscopy for cell behavioural examination and manipulation”, Fondazione Patrimonio Ca’ Granda “Liver Bible” PR-0361. DP was supported by Ricerca corrente Fondazione IRCCS Ca’ Granda Ospedale Maggiore Policlinico, CV PREVITAL “Strategie di prevenzione primaria nella popolazione Italiana” Ministero della Salute, and Associazione Italiana per la Prevenzione dell’Epatite Virale (COPEV).

Genetic modifiers for COVID-19 related illness (BeLCovid_1) was supported by the “Fonds Erasme”. The Host genetics and immune response in SARS-Cov-2 infection (BelCovid_2) study was supported by grants from Fondation Léon Fredericq and from Fonds de la Recherche Scientifique (FNRS).

The INMUNGEN-CoV2 study was funded by the Consejo Superior de Investigaciones Científicas.

KUL is supported by the German Research Foundation (LU 1944/3-1)

SweCovid is funded by the SciLifeLab/KAW national COVID-19 research program project grant to Michael Hultström (KAW 2020.0182) and the Swedish Research Council to Robert Frithiof (2014-02569 and 2014-07606). HZ is supported by Jeansson Stiftelser, Magnus Bergvalls Stiftelse.

The COMRI cohort is funded by Technical University of Munich, Munich, Germany. Genotyping for the COMRI cohort was performed and funded by the Genotyping Laboratory of Institute for Molecular Medicine Finland FIMM Technology Centre, University of Helsinki, Helsinki, Finland.

These funding agencies had no role in the design, implementation or interpretation of this study.

## Declaration of interests

JBR has served as an advisor to GlaxoSmithKline and Deerfield Capital. DP has served as an advisory board, and has received travel/research grants, speaking and teaching fees for Macopharma, Ortho Clinical Diagnostics, Grifols, Terumo, Immucor, Diamed, and Diatech Pharmacogenetics. THK has served an advisor to Novartis, Gilead, Intercept and Engitix. LV declares following; speaking fees: MSD, Gilead, AlfaSigma, AbbVie, Consulting: Gilead, Pfizer, Astra Zeneca, Novo Nordisk, Intercept pharmaceuticals, Diatech Pharmacogenetics, IONIS; Research grants: Gilead. All other authors declare that there are no conflicts of interest.

## Contributors

Conception and design: TN, GBL, BNJ, FG, RF, MRG, KUL, MB, SR, MEAR, ECS, THK, LV, HZ, JBR and AG. Formal analysis: TN, SP, FD, MC, GBL, DMM, BNJ, YB, MN, DE, MMB, KUL, MEAR, LV, HZ, BR and AG. Data curation: TN, FD, MC, GBL, DMM, BNJ, YB, MN, DE, MMB, SR, SA, LR, FF, CDS, FG, IFC, JCH, RF, RA, ACP, LB, JRH, IM, AR, KUL, MB, ECS, JBR and AG. Interpretation of data: TN, GBL, BNJ, SR, RF, MRG, IM, KUL, MEAR, LV, HZ, BR, and AG. Funding acquisition: DMM, SA, FF, CDS, DP, DB, FG, GD, JCH, RF, SD, MRG, JRH, IM, AR, KUL, MB, SR, MEAR, ECS, THK, JBR, and AG. Investigation: TN, GBL, DMM, BNJ, YB, RF, IM, KUL, MEAR, BR and AG. Methodology: TN, GBL, MMB, MEAR, HZ, JBR and AG. Project administration: TN, FD, DMM, SR, CDS, DP, DB, FG, GD, JCH, JB, JRH, IM, KUL, SR, ECS, AF, THK, LV, JBR and AG. Resources: FG, GD, MRG, IM, SR, MEAR, JBR and AG. Supervision: DMM, BNJ, FG, MRG, IM, KUL, SR, MEAR, JBR and AG. Validation: TN, SP, FD, DE, AK, KK, and AG. Visualization: TN and AG. Writing—original draft: TN, JBR and AG. Writing—review and editing: TN, GBL, DMM, BNJ, AP, SR, IFC, JCH, RF, KK, SD, RA, LB, JRH, IM, AR, AMP, KUL, MEAR, THK, LV, HZ, JBR and AG. All authors were involved in further drafts of the manuscript and revised it critically for content. All authors gave final approval of the version to be published. The corresponding authors attest that all listed authors meet authorship criteria and that no others meeting the criteria have been omitted.

## Ethical approval

All institutions contributing cohorts to the COVID-19 Host Genetics Initiative received ethics approval from their respective research ethics review boards. Genetic modifiers for COVID-19 related illness (BelCovid_1) was approved by the Erasme Ethics committee (protocol P2020_209). Host genetics and immune response in SARS-Cov-2 infection (BelCovid_2) was approved by the ethics committee of Liege University Hospital (approval number 2020-242). The BoSCO study was approved by Ethics Committee of the Medical Faculty of the University of Bonn. BQC19 received ethical approval from the JGH research ethics board (2020-2137). The BRACOVID study has been approved by the Hospital das Clinicas, Sao Paulo University Medical School and by Brazilian National IRB, CONEP. COMRI and the COVID-19 biobank of the Faculty of Medicine at Technical University Munich received ethical approval from the local research ethics board (TUM 217/20, TUM 221/20S, TUM 440/20S). San Sebastian Hospital and Basque Biobank (COVID19-Host(a)ge_1) was approved by the Euskadi Ethics Committee on April 6, 2020 (approval number PI2020064). The study in Hospital Universitario Valle Hebron and CIberehd del Instituto Carlos III. Barcelona (COVID19-Host(a)ge_2) was approved by Vall d’Hebron Ethical Committee. COVID GWAs, Premed COVID-19 (COVID19-Host(a)ge_3) was approved by COVID GWAs (ethics id: 0886-N-20) and Premed Covid (ethics id: 1954-N-20). Genetics against coronavirus (GENIUS), Humanitas University (COVID19-Host(a)ge_4) was approved by the ethic committee (approval number reference number 316/20). FoGS was approved by the ethics committee (approval number 342_2020). The GEN-COVID is a multicentre academic observational study was approved by the IRB of each participating centre. The INMUNGEN-CoV2 study was reviewed and approved by the Ethical Committee of the Hospital Clinic of Barcelona (CEIm number: Reg.HCB/2020/0357). NorCoV2 was approved by the Regional Committee for Medical and Health Research Ethics in South-Eastern Norway (project no. 132550). SPGRX was reviewed and approved by the Valladolid Ethics Committee (PI-201716) and the Granada Ethics Committee (no number given) on March 24th, 2020 and April 13th, 2020, respectively. SweCovid was approved by the National Ethical Review Agency (EPM; 2020-01623). UK Biobank was approved by the Northwest Multi-Centre Research Ethics Committee and informed consent was obtained from all participants prior to participation. This study was conducted under project ID 27449. FinnGen was approved by HUS coordinating Ethics committee. The Columbia University Biobank was approved by the Columbia University IRB.

## Data sharing

The harmonized individual-level data of some participating cohorts from Belgium (BeLCovid_2), Brazil (BRACOVID), Italy (COVID19-Host(a)ge_4, GEN-COVID), Spain (COVID19-Host(a)ge_1,2,3, INMUNGEN-CoV2, SPGRX), and Sweden (SweCovid) is under preparation to be deposited at the European Genome-phenome Archive (EGA).

Regarding the data from genetic modifiers for COVID-19 related illness (BelCovid_1), individual level data were acquired and shared with FIMM during the sanitary crisis under an emergency consent and an ethical approval which were specific to this particular project and do not cover deposition to public repositories. Upon contact with Françoise Wilkin, Isabelle Migeotte, or Guillaume Smits, an institutional data transfer agreement can be established and data shared if the aims of data use are covered by ethical approval and patient consent. The procedure will involve an update to the ethical approval, as well as review by legal departments at both institutions and the process will typically take 2-4 months from initial contact.

Regarding the BoSCO study, individual-level genotype and clinical data for purpose of this study were shared with FIMM under a legal, bilateral agreement and were specific to this particular project. Current participant consents and privacy regulations prohibit deposition of individual level data to public repositories. Upon contact with Kerstin Ludwig or Markus M. Nöthen, an institutional data transfer agreement can be established and data shared if the aims of data use is covered by ethical approvals and patient consent. The procedure will involve review by legal departments at both institutions and the process will typically take about 2 months from initial contact.

The BQC19 is an Open Science biobank. Instructions on how to access data for individuals from the BQC19 at the Jewish General Hospital site are available here: https://www.mcgill.ca/genepi/mcg-covid-19-biobank. Instructions on how to access data from other sites of the BQC19 are available here: https://www.bqc19.ca/en/access-data-samples. For the COMRI cohort, data protection legislation does not allow for deposition of individual level data in public repositories. Upon direct contact with Prof Ulrike Protzer (, genetic data) and Dr Christoph Spinner, an institutional data transfer agreement can be established and data will be shared if the aims of data use are covered by ethical approvals and patient consent. The procedure will involve an update to the ethical approval as well as review by legal departments at both institutions and the process will typically take 2-3 months from initial contact.

Regarding the Fondazione IRCCS Milan data (FOGS study), institutional data privacy regulations prohibit deposition of individual level data to public repositories without a specific consent. Participant written consent also does not cover public sharing of data for use for unknown purposes. Upon contact with professor Luca Valenti an institutional data transfer agreement can be established and data shared if the aims of data use are covered by ethical approvals and patient consent. The procedure will involve the request for an amendment to the ethical approvals, as well as review by legal departments at both institutions and the process will typically take 1-2 months from initial contact.

Regarding Norwegian data (NorCoV2), institutional data privacy regulations prohibit deposition of individual level data to public repositories. Participant written consent also does not cover public sharing of data for use for unknown purposes. Upon contact with professor Tom H Karlsen or professor Johannes R. Hov an institutional data transfer agreement can be established and data shared if the aims of data use is covered by ethical approvals and patient consent. The procedure will involve an update to the ethical approvals, as well as review by legal departments at both institutions and the process will typically take 1-2 months from initial contact.

The genetic and phenotype datasets from UK Biobank are available via the UK Biobank data access process (see http://www.ukbiobank.ac.uk/register-apply/).

## Code availability

All code for data management and analysis is archived online at https://github.com/tomoconaka/COVID19-chr3 for review and reuse.

## Reference

1 McKee M, Stuckler D. If the world fails to protect the economy, COVID-19 will damage health not just now but also in the future. Nat. Med. 2020; 26: 640–2.

2 Buitrago-Garcia D, Egli-Gany D, Counotte MJ, et al. Occurrence and transmission potential of asymptomatic and presymptomatic SARSCoV-2 infections: A living systematic review and meta-analysis. PLoS Med. 2020; 17: e1003346.

3 Dooling K, Marin M, Wallace M, et al. The Advisory Committee on Immunization Practices’ Updated Interim Recommendation for Allocation of COVID-19 Vaccine — United States, December 2020. MMWR Morb Mortal Wkly Rep 2021; 69: 1657–60.

4 O’Driscoll M, Dos Santos GR, Wang L, et al. Age-specific mortality and immunity patterns of SARS-CoV-2. Nature 2020; 590: 140–5.

5 Williamson EJ, Walker AJ, Bhaskaran K, et al. Factors associated with COVID-19-related death using OpenSAFELY. Nature 2020; 584: 430–6.

6 Chaudhry R, Dranitsaris G, Mubashir T, Bartoszko J, Riazi S. A country level analysis measuring the impact of government actions, country preparedness and socioeconomic factors on COVID-19 mortality and related health outcomes. EClinicalMedicine 2020; 25: 100464.

7 Baric RS. Emergence of a Highly Fit SARS-CoV-2 Variant. N Engl J Med 2020; 383: 2684–6.

8 Van Der Kolk DM, De Bock GH, Leegte BK, et al. Penetrance of breast cancer, ovarian cancer and contralateral breast cancer in BRCA1 and BRCA2 families: High cancer incidence at older age. Breast Cancer Res Treat 2010; 124: 643–51.

9 Nordestgaard BG, Chapman MJ, Humphries SE, et al. Familial hypercholesterolaemia is underdiagnosed and undertreated in the general population: guidance for clinicians to prevent coronary heart disease: Consensus Statement of the European Atherosclerosis Society. Eur Heart J 2013; 34: 3478–90.

10 Biobank U, Feng Y-CA, Ge T, et al. Findings and insights from the genetic investigation of age of first reported occurrence for complex disorders in the UK Biobank and FinnGen. medRxiv 2020; : 2020.11.20.20234302.

11 Ellinghaus D, Degenhardt F, Bujanda L, et al. Genomewide Association Study of Severe Covid-19 with Respiratory Failure. N Engl J Med 2020; : EJMoa2020283.

12 Pairo-Castineira E, Clohisey S, Klaric L, et al. Genetic mechanisms of critical illness in Covid-19. Nature 2020; : 1–1.

13 The COVID-19 Host Genetics Initiative, a global initiative to elucidate the role of host genetic factors in susceptibility and severity of the SARS-CoV-2 virus pandemic. Eur J Hum Genet 2020; 28: 715–8.

14 Zeberg H, Pääbo S. The major genetic risk factor for severe COVID-19 is inherited from Neanderthals. Nature 2020; 587: 610–2.

15 Karczewski KJ, Francioli LC, Tiao G, et al. The mutational constraint spectrum quantified from variation in 141,456 humans. Nature 2020; 581: 434–43.

16 COVID-19 Host Genetics Analysis Plan v1.1 - Google Docs. https://docs.google.com/document/d/16ethjgi4MzlQeO0KAW_yDYyUHdB9kKbtfuGW4XYVKQg/edit#heading=h.yvzvdf3jx6u9 (accessed Feb 7, 2021).

17 Auton A, Abecasis GR, Altshuler DM, et al. A global reference for human genetic variation. Nature. 2015; 526: 68–74.

18 Altshuler DM, Gibbs RA, Peltonen L, et al. Integrating common and rare genetic variation in diverse human populations. Nature 2010; 467: 52–8.

19 Del Valle DM, Kim-Schulze S, Huang H-H, et al. An inflammatory cytokine signature predicts COVID-19 severity and survival. Nat Med 2020; : 1–8.

20 Merrill JT, Erkan D, Winakur J, James JA. Emerging evidence of a COVID-19 thrombotic syndrome has treatment implications. Nat. Rev. Rheumatol. 2020; 16: 581–9.

21 Higuera-de la Tijera F, Servín-Caamaño A, Reyes-Herrera D, et al. Impact of liver enzymes on SARS-CoV-2 infection and the severity of clinical course of COVID-19. Liver Res 2021;published online Jan 12. DOI:10.1016/j.livres.2021.01.001.

22 Vafadar Moradi E, Teimouri A, Rezaee R, et al. Increased age, neutrophil-to-lymphocyte ratio (NLR) and white blood cells count are associated with higher COVID-19 mortality. Am J Emerg Med 2021; 40: 11–4.

23 Yan L, Zhang H-T, Goncalves J, et al. An interpretable mortality prediction model for COVID-19 patients. Nat Mach Intell 2020; 2: 283–8.

24 Données COVID-19 par âge et sexe au Québec | INSPQ. https://www.inspq.qc.ca/covid-19/donnees/age-sexe (accessed Feb 14, 2021).

25 Signorelli C, Odone A. Age-specific COVID-19 case-fatality rate: no evidence of changes over time. Int. J. Public Health. 2020; 65: 1435–6.

26 Certain Medical Conditions and Risk for Severe COVID-19 Illness | CDC. https://www.cdc.gov/coronavirus/2019-ncov/need-extra-precautions/people-with-medical-conditions.html (accessed Jan 29, 2021).

27 Purcell S, Neale B, Todd-Brown K, et al. PLINK: A tool set for whole-genome association and population-based linkage analyses. Am J Hum Genet 2007; 81: 559–75.

28 Vuille-Dit-Bille Rn, Camargo SM, Emmenegger L, et al. Human intestine luminal ACE2 and amino acid transporter expression increased by ACE-inhibitors. Amino Acids 2015; 47: 693–705.

29 Yao Y, Ye F, Li K, et al. Genome and epigenome editing identify CCR9 and SLC6A20 as target genes at the 3p21.31 locus associated with severe COVID-19. Signal Transduct Target Ther 2021; 6: 85.

30 Smieszek SP, Polymeropoulos MH. Role of FYVE and Coiled-Coil Domain Autophagy Adaptor 1 in severity of COVID-19 1 infection 2. medRxiv 2021; : 2021.01.22.21250070.

31 Payne DJ, Dalal S, Leach R, et al. The CXCR6/CXCL16 axis links inflamm-aging to disease severity in COVID-19 patients 2 3. DOI:10.1101/2021.01.25.428125.

32 Khalil BA, Elemam NM, Maghazachi AA. Chemokines and chemokine receptors during COVID-19 infection. Comput. Struct. Biotechnol. J. 2021; 19: 976–88.

33 Chua RL, Lukassen S, Trump S, et al. COVID-19 severity correlates with airway epithelium–immune cell interactions identified by single-cell analysis. Nat Biotechnol 2020; 38: 970–9.

34 Liao M, Liu Y, Yuan J, et al. Single-cell landscape of bronchoalveolar immune cells in patients with COVID-19. Nat Med 2020; 26: 842–4.

35 Fry A, Littlejohns TJ, Sudlow C, et al. Comparison of Sociodemographic and Health-Related Characteristics of UK Biobank Participants With Those of the General Population. Am J Epidemiol 2017; 186: 1026–34.

36 Griffith GJ, Morris TT, Tudball MJ, et al. Collider bias undermines our understanding of COVID-19 disease risk and severity. Nat Commun 2020; 11: 1–12.

