## Supplementary Document and Figures for "Age-dependent impact of the major common genetic risk factor for COVID-19 on severity and mortality"

### Supplementary cohort descriptions

#### **Individual study information in COVID-19 HGI**

This section contains the individual study information of all studies with available individual-level data (referred to as COVID-19 HGI cohorts). The cohorts are listed alphabetically according to the abbreviated name of each study. **Supplementary Table 4** summarizes study-specific demographic and clinical data, as well as the missingness rates for each variable.

##### **Genetic modifiers for COVID-19 related illness (BeLCovid_1)**

Isabelle Migeotte, Youssef Bouysran, Adeline Busson, Xavier Peyrassol, Françoise Wilkin, Bruno Pichon, Guillaume Smits, Isabelle Vandernoot, Jean-Christophe Goffard, Nicky Tiembe

The study was designed within the frame of the COVID-19 host genetics initiative (HGI) and aimed at participating to the search for genetic determinants of COVID-19 severity and outcome through GWAS analysis of hospitalized patients. All patients present in Hôpital Erasme (Brussels) between April and June 2020 and displaying a positive SARS-Cov2 PCR test were recruited. DNA samples and corresponding anonymized demographic and clinical data were sent to Finland Institute for Molecular Medicine where genotyping was performed. The study was reviewed and approved by the Erasme Ethics committee (protocol P2020_209).

**Clinical data**

Individual level patient data encompassing demographic data, diagnoses, test results, and treatments were extracted from the WHO questionnaires and completed to match the common template defined by the HGI. Data were obtained by medical chart review performed by trained clinicians. For biochemical and haematological tests, results were obtained by accessing the electronic medical record system database.

**Genotype data**

DNA was extracted from blood collected on EDTA tubes through treatment by Proteinase K followed by extraction using Maxwell RSC Blood DNA kit (Promega). Genotyping and imputation were performed by the Genotyping laboratory of Institute for Molecular Medicine Finland FIMM Technology Centre, University of Helsinki. (Details are described in genotyping QC and imputation at FIMM section).

##### **Host genetics and immune response in SARS-Cov-2 infection (BelCovid_2)**

Souad Rahmouni, Gilles Darcis, Michel Georges, Michel Moutschen, Benoit Misset, Julien Guiot, Julien Guntz, Gilles Parzibut, Christelle Meuris, Marie Thys, Jessica Jacques, Philippe Léonard, Frederic Frippiat, Jean-Baptiste Giot, Anne-Sophie Sauvage, Christian Von Frenckell, Bernard Lambermont, Olivier Malaise, Christophe Bovy, Antoine Bouquegneau

Patients admitted to Liège University Hospital (CHU of Liège) and to the clinic CHC-MontLégia (Liege, Belgium) and tested positive for SARS-CoV-2 were invited to participate in the study. Additionally, hospital asymptomatic staff members (including healthcare workers and administrative agents) enrolled into a prospective study implemented as an active monitoring protocol for COVID-19 aiming at detecting SARS-CoV-2 infection at the earliest stage were invited to participate in the study when tested positive for the SARS-CoV-2. Samples were collected between March 25 and December 31, 2020. Demographics, clinical data and samples were collected after the study participant had acknowledged that they had understood the study protocol and signed the informed consent. The protocol was reviewed and approved by the ethics committee of Liege University Hospital (approval number 2020-242). All collected samples and data were made available to us through the Liege Biobank (BHUL: [www.gigabiotheque.uliege.be](http://www.gigabiotheque.uliege.be)).

**Clinical data**

Demographics, clinical, diagnosis and testing data were collected by the clinicians in charge of the patients and were stored in the internal clinical databases of the participating clinical centers. Access was granted to us through the Liege Biobank (BHUL).

**Genotype data**

DNA was extracted from whole blood. Genotyping and imputation were performed by the Genotyping laboratory of Institute for Molecular Medicine Finland FIMM Technology Centre, University of Helsinki. (Details are described in genotyping QC and imputation at FIMM section).

##### **Bonn Study of COVID19 genetics (BosCO)**

Kerstin U. Ludwig, Selina Rolker, Markus M. Nöthen, Julia Fazaal, Verena Keitel, Björn Jensen, Torsten Feldt, Ingo Kurth, Nikolaus Marx, Michael Dreher, Isabell Pink, Markus Cornberg, Thomas Illig, Clara Lehmann, Philipp Schommers, Max Augustin, Jan Rybniker, Lisa Knopp, Thomas Eggermann, Sonja Volland, Alexander Popov, Janine Altmüller, Julia Schröder, Carlo Maj

The BoSCO study was established in May 2020. It recruits probands via two different arms, i.e., population-based and through collaborations with other university hospitals, with the major aim to contribute to the COVID19-HGI. Ethic approval has been obtained at each clinical site including Ethics Committee of the Medical Faculty of the University of Bonn, and informed consent was provided by each participant. For this specific study, a subset of patients was selected, including only those probands who had been hospitalized due to COVID19.

**Clinical data**

For each patient, the respective clinical data were retrieved from the respective medical institutions, or from the patients themselves through questionnaires.

**Genotype data**

DNA was extracted either from saliva or from blood, both using standard procedures. Genotyping was performed using the Illumina GSA v3.0 array, at the Genomics Department of Life&Brain Center, Bonn, Germany. Subsequent quality control (including removal of relatives or samples with low call rate) was performed in accordance with the data analysis plan provided by the COVID19-HGI^1^. The minor allele frequency (MAF) and imputation score for rs10490770 was 0.12 and 0.97, respectively.

##### **Biobanque Quebec COVID19 (BQC19)**

Tomoko Nakanishi, Guillaume Butler-Laporte, Vincenzo Forgetta, David R. Morrison, Biswarup Ghosh, Laetitia Laurent, Alexandre Belisle, Rui Li, Danielle Henry, Tala Abdullah, Olumide Adeleye, Noor Mamlouk, Nofar Kimchi, Zaman Afrasiabi, Nardin Rezk, Branka Vulesevic, Meriem Bouab, Charlotte Guzman, Louis Petitjean, Chris Tselios, Xiaoqing Xue, Erwin Schurr, Jonathan Afilalo, Marc Afilalo, Maureen Oliveira, Bluma Brenner, Pierre Lepage, Jiannis Ragoussis, Daniel Auld, G. Mark Lathrop, Vincent Mooser, J. Brent Richards

The Biobanque Québec COVID-19 (www.BQC19.ca) is a provincial biobank that prospectively enrolls patients with suspected COVID-19, or COVID-19 confirmed through SARS-CoV-2 PCR testing. For this study, we used results from patients with available genotype data recruited at the Jewish General Hospital (JGH), a university affiliated tertiary care center serving a multiethnic population in Montréal, Québec, Canada. BQC19 received ethical approval from the JGH research ethics board (2020-2137).

**Clinical data**

Individual level patient data encompassing demographic data, test results, diagnoses, and treatments were obtained by medical chart review or patient interview performed by trained clinicians or research coordinators. For biochemical and haematological tests, results were obtained by accessing the electronic medical record system database directly, rather than through chart reviews.

**Genotype data**

SNP genotyping was conducted using the Axiom™ Precision Medicine Research Array from Applied Biosystems™^2^. We used 17 plates array of the 96 well plate format. There were 92 studied samples per plate and 4 controls; 1 time NA24385, 2 times NA12878 and one negative control. All sample preparation was done using sample handlers. Normalization of 200 ng of DNA extracted from whole blood of each sample was done using a Perkin Elmer JANUS Liquid Handler. After that, molecular process was entirely done using an Axiom™ NIMBUS (Hamilton Robotics). Staining and scanning of the plate array was done using an Applied Biosystems™ GeneTitan™ MC instrument. All the samples included in the study were grouped together, using the Axiom Analysis Suite 5.1.1 software, to perform the “Best Practice Workflow” analysis using high quality call rate parameters: “Axiom_PMRA.r3” library and threshold configuration “Human.v5” with minimum call rate set at 97.0%.

Marker quality control tests were performed on a subset of ancestrally homogeneous participants genotyped at 861,229 markers, who were determined via comparison to 2,504 individuals across 5 super populations from 1000 Genomes phase 3^3^. We performed batch effect quality control, replicate discordance check, and finally variants with low allele frequency (MAF<0.001), low genotyping call rate (<98%), and with departure from Hardy-Weinberg equilibrium (HWE) (p value<1x10^-6^) were removed. This left 536,222 high quality genotype markers.

For sample quality control, we used the 536,222 genetic markers above and further filtered them using the following additional criteria: MAF>0.01, marker-wise missingness <0.01, single nucleotide substitutions with single character allele-codes (A, C, G, or T) (PLINK --snps-only ‘just-acgt’ option), and excluded variants within high linkage disequilibrium (LD) region. The above filtering resulted in a total of 407,158 genetic markers, which were used to determine low quality samples. We removed individuals with extreme values in heterozygosity and missingness. These outliers are defined as high genotype missing rate (overall missing genotype rate >0.03) or high/low heterozygosity rate on autosomal chromosomes (>0.22 or <0.17, which are equivalent to +3SD in AFR population and -3SD in EAS population, respectively, in our cohort). We then determined sex chromosome composition by estimating heterozygosity of X chromosome markers using PLINK (--check-sex 0.45 0.8). Since distribution of chromosome X heterozygosity estimates (F estimates) showed a gap between 0.45 and 0.8, we obtained sex chromosome numbers and compared these to self-reported gender. There were two individuals with discordant self-reported gender and genetic sex, and thus removed for the imputation step. The sample quality control process left 1,334 samples, which were used for genotype imputation. We performed genotype phasing and imputation on the 1,334 BQC19 participants using the TOPMed reference panel^4,5^ at the University of Michigan Imputation Service^6^. The MAF and imputation score for rs10490770 was 0.10 and 0.96, respectively.

##### **Genetic determinants of COVID-19 complications in the Brazilian population (BRACOVID)**

Jose E Krieger, Alexandre C Pereira, Cinthia E Jannes, Isabella Ramos Lima, Mauricio Teruo Tada, Mariliza Velho, Emanuelle Marques, Karina Valino

Individuals with positive SARS-Cov-2 virus identification or with serology were collected from hospitalized patients at the Hospital das Clinicas, Sao Paulo University Medical School from April 2020. The study has been approved by the Hospital das Clinicas, Sao Paulo University Medical School and by Brazilian National IRB, CONEP.

**Clinical data**

Individual level patient data encompassing demographic data, test results, diagnoses, and treatments were obtained by medical chart review or patient interview performed by trained clinicians or research coordinators.

**Genotype data**

DNA was extracted from whole blood. Genotyping was performed with the Axiom_PMRA.r3 array. Imputation was performed by the Genotyping laboratory of Institute for Molecular Medicine Finland FIMM Technology Centre, University of Helsinki. (Details are described in genotyping QC and imputation at FIMM section). The MAF and imputation score for rs10490770 was 0.06 and 0.96, respectively.

##### **COVID-19 Cohort Study of the University Medical Center of the Technical University Munich (COMRI)**

Eva C. Schulte, Christoph D. Spinner, Ulrike Protzer, Clemens-Martin Wendtner, Christof Winter, Johanna Erber

COMRI is an observational study prospectively enrolling individuals with suspected or confirmed COVID-19 and treated at the University Medical Center of the Technical University Munich. Biosamples are deposited in the COVID-19 biobank of the Faculty of Medicine of the Technical University Munich. For this study, we used results from patients with PCR-confirmed SARS-CoV-2 infection recruited between March 1 and August 31, 2020, and available genome-wide genotyping data. COMRI and the COVID-19 biobank of the Faculty of Medicine at Technical University Munich received ethical approval from the local research ethics board (TUM 217/20, TUM 221/20S, TUM 440/20S).

**Clinical data**

Individual level patient data encompassing demographic data, test results, diagnoses, and treatments were obtained by medical chart review or patient interview performed by trained clinicians or research coordinators. For biochemical and haematological tests, results were curated manually via the electronic medical record system database.

**Genotype data**

DNA was extracted from whole blood. Genotyping and imputation were performed by the Genotyping laboratory of Institute for Molecular Medicine Finland FIMM Technology Centre, University of Helsinki. (Details are described in genotyping QC and imputation at FIMM section).

##### **San Sebastian Hospital and Basque Biobank (COVID19-Host(a)ge_1)**

Luis Bujanda, Jesus Banales, Beatriz Nafria-Jimenez, Adolfo Garrido Chercoles, Koldo Garcia-Etxebarria, Pedro M. Rodrigues, Laura Izquierdo, Mauro D'Amato, Eunate Arana, Josune Goikoetxea, Natale Imaz Ayo, Maider Intxausti, Cristina Sancho, Pedro P. España, Eloisa Urrechaga, María A. Gutiérrez-Stampa

Patients with positive SARS-CoV-2 and severe pneumonia who were admitted to the University Hospitals of Donostia, Cruces, Galdakao and Basurto (Basque Country) were included. The patients gave their consent to study the genetic factors that predispose to infection and to develop serious complications. Samples were collected between April and December, 2020. The protocol was reviewed and approved by the Euskadi Ethics Committee on April 6, 2020 (approval number PI2020064).

**Clinical data**

Individual level patient data encompassing demographic data, test results, diagnoses, and treatments were obtained by medical chart review or patient interview performed by trained clinicians or research coordinators.

**Genotype data**

DNA extraction from blood samples, genotyping and imputation have been done at and by University of Kiel, Department of Molecular Medicine. (Details are described in genotype QC and imputation at Kiel section).

##### **Hospital Universitario Valle Hebron and CIberehd del Instituto Carlos III. Barcelona (COVID19-Host(a)ge_2)**

Maria Butti, Adriana Palom, Luisa Roade

Samples were collected for the COVID-19 HGI including patients who tested positive in Vall d'Hebron Hospital laboratory. Anonymous data collection at individual level was approved by Vall d'Hebron Ethical Committee.

**Clinical data**

Individual level data including demographic, laboratory and clinical data of admitted patients were obtained by medical chart review performed by trained clinicians.

**Genotype data**

DNA extraction from blood samples, genotyping and imputation have been done at and by University of Kiel, Department of Molecular Medicine. (Details are described in genotype QC and imputation at Kiel section).

##### **COVID GWAs, Premed COVID-19 (COVID19-Host(a)ge_3)**

Manuel Romero-Gómez, Douglas Maya-Miles, Enrique Calderon, Adolfo de Salazar, Jose Hernández Quero, Natalia Chueca, Trinidad Gonzalez Cejudo, Carmen de la Horra, Francisco J. Medrano, Javier Ampuero, Juan Delgado, Juan M. Guerrero, Rocío Gallego-Durán, Rubén Morilla, Vicente Friaza, Ximo Dopazo

Samples come from two studies funded by the Andalusian Government: COVID GWAs (ethics id: 0886-N-20) and Premed Covid (ethics id: 1954-N-20) to search for genetic host and viral factors associated to COVID-19 severity and come mostly from patients from the Hospital Universitario Virgen del Rocío and the Hospital Universitario San Cecilio.

**Clinical data**

Individual level patient data encompassing demographic data, diagnoses, test results, and treatments were obtained by accessing the Andalusian electronic medical record system database. Data were obtained by medical chart review performed by trained clinicians.

**Genotype data**

DNA extraction from blood samples, genotyping and imputation have been done at and by University of Kiel, Department of Molecular Medicine. (Details are described in genotype QC and imputation at Kiel section).

##### **Genetics against coronavirus (GENIUS), Humanitas University (COVID19-Host(a)ge_4)**

Rosanna Asselta, Stefano Duga, Alberto Mantovani, Alessandro Protti, Alessio Aghemo, Ana Lleo, Antonio Voza, Claudio Cappadona, Elena Azzolini, Elvezia Maria Paraboschi, Ilaria My, Massimo Castoldi, Maurizio Cecconi, Paolo Tentorio, Salvatore Badalamenti, Sara Bombace, Valeria Rimoldi, Chiara Masetti, Francesca Colapietro, Simone Solano, Viviana Barbieri, The Humanitas Gavazzeni COVID-19 Task Force, Humanitas COVID-19 Task Force

SARS-CoV-2 positive patients with severe lung affection defined by hospitalization and respiratory failure at the Humanitas clinical and research center, whose blood samples were collected between March and April 2020, were recruited for the study after the submission of informed consent. The aims of the project were/are to identify the main common genetic determinants of severe COVID-19 by conducting a GWAS, to identify rare genetic variants of severe COVID-19 by conducting whole-exome sequencing, and to provide a coordination to manage the collaboration with other research consortia in the field, e.g. the "COVID-19 Host Genetic Initiative" to develop a genetic risk score to stratify disease risk, to assess the impact of genetic risk variants on disease outcome and finally to examine the possible role of rare genetic variants in determining the predisposition to severe CODIV-19. Genotyping have been performed by the Illumina GlobalScreeningArray-24 v3.0. The protocol was reviewed and approved by the ethic committee (approval number reference number 316/20).

**Clinical data**

Patient data comprised demographic data, test results, diagnoses, and treatments. These data were collected by the clinicians in charge of the patients and were stored in the internal clinical electronic database of the participating clinical centers.

**Genotype data**

DNA extraction from blood samples, genotyping and imputation have been done at and by University of Kiel, Department of Molecular Medicine. (Details are described in genotype QC and imputation at Kiel section).

##### **Fondazione IRCCS Ca' Granda Ospedale Maggiore Policlinico, Università degli Studi di Milano (FoGS)**

Daniele Prati, Luca Valenti, Guido Baselli, Luigi Santoro, Serena Pelusi, Filippo Martinelli-Boneschi, Luigia Scudeller, Silvano Bosari, Alberto Zanella, Giacomo Grasselli, Antonio Pesenti, Valter Monzani, Francesco Blasi, Andrea Gori, Alessandra Bandera, Flora Peyvandi, Roberta Gualtierotti, Giorgio Costantino, Anna Ludovica Fracanzani, Marina Baldini, Maria Carrabba, Leonardo Terranova, Ferruccio Ceriotti, Monica Miozzo, Nicola Montano, Stefano Aliberti, Cinzia Paccapelo, Alessandro Cherubini, Cristiana Bianco, Francesco Malvestiti, Giuseppe Lamorte, Cinzia Hu, Melissa Tomasi, Angela Lombardi, Rossana Carpani

SARS-CoV-2 positive patients with severe lung affection defined by hospitalization and respiratory failure at the Fondazione IRCCS Ca’ Granda, whose blood samples were collected between March and April 2020, were recruited for the study after the submission of informed consent. The specific aims of the project are to identify the main common genetic determinants of severe COVID-19 by conducting a GWAS, to establish a database and a biorepository of samples from donors which have reported an asymptomatic/subclinical infection, to provide a genetic characterization of all patients managed by the Fondazione and to provide a coordination to manage the collaboration with other research consortia in the field e.g. the “An anonymized GWAS to urgently query host genetic predisposition to severe COVID-19 lung disease” and the “COVID-19 Host Genetic Initiative” in order to develop a genetic risk score to stratify disease risk, to assess the impact of genetic risk variants on disease outcome and finally to examine the possible role of rare genetic variants in determining the predisposition to severe COVID-19. The protocol was reviewed and approved by the ethics committee (approval number 342_2020).

**Clinical data**

Individual level patient data encompassing demographic data, test results, diagnoses, and treatments were collected by the clinicians in charge of the patients and were stored in the internal clinical databases.

**Genotype data**

DNA extraction from blood samples, genotyping and Imputation have been done at and by University of Kiel, Department of Molecular Medicine. (Details are described in genotype QC and imputation at Kiel section).

##### **GEN-COVID**

Alessandra Renieri, Francesca Mari, Chiara Fallerini, Sergio Daga, Elisa Benetti, Margherita Baldassarri, Francesca Fava, Elisa Frullanti, Floriana Valentino, Gabriella Doddato, Annarita Giliberti, Rossella Tita, Sara Amitrano, Mirella Bruttini, Susanna Croci, Ilaria Meloni, Maria Antonietta Mencarelli, Caterina Lo Rizzo, Anna Maria Pinto, Giada Beligni, Andrea Tommasi, Laura Di Sarno, Maria Palmieri, Miriam Lucia Carriero, Diana Alaverdian, Nicola Iuso, Gabriele Inchingolo, Nicola Picchiotti, Maurizio Sanarico, Marco Gori, Simone Furini, Stefania Mantovani, Raffaele Bruno, Marco Vecchia, Serena Ludovisi, Mario Umberto Mondelli, Francesco Castelli, Eugenia Quiros-Roldan, Melania Degli Antoni, Isabella Zanella, Massimo Vaghi, Stefano Rusconi, Matteo Siano, Francesca Montagnani, Massimiliano Fabbiani, Barbara Rossetti, Giacomo Zanelli, Elena Bargagli, Laura Bergantini, Miriana D’Alessandro, Paolo Cameli, David Bennet, Federico Anedda, Simona Marcantonio, Sabino Scolletta, Federico Franchi, Maria Antonietta Mazzei, Susanna Guerrini, Edoardo Conticini, Luca Cantarini, Bruno Frediani, Danilo Tacconi, Chiara Spertilli, Marco Feri, Alice Donati, Raffaele Scala, Luca Guidelli, Genni Spargi, Marta Corridi, Cesira Nencioni, Leonardo Croci, Maria Bandini, Gian Piero Caldarelli, Maurizio Spagnesi, Paolo Piacentini, Elena Desanctis, Silvia Cappelli, Anna Canaccini, Agnese Verzuri, Valentina Anemoli, Agostino Ognibene, Alessandro Pancrazi, Maria Lorubbio, Antonella D’Arminio Monforte, Esther Merlini, Federica Gaia Miraglia, Massimo Girardis, Sophie Venturelli, Stefano Busani, Andrea Cossarizza, Andrea Antinori, Alessandra Vergori, Arianna Emiliozzi, Arianna Gabrieli, Agostino Riva, Daniela Francisci, Elisabetta Schiaroli, Francesco Paciosi, Pier Giorgio Scotton, Francesca Andretta, Sandro Panese, Renzo Scaggiante, Francesca Gatti, Saverio Giuseppe Parisi, Stefano Baratti, Matteo Della Monica, Carmelo Piscopo, Mario Capasso, Roberta Russo, Immacolata Andolfo, Achille Iolascon, Giuseppe Fiorentino, Massimo Carella, Marco Castori, Giuseppe Merla, Gabriella Maria Squeo, Filippo Aucella, Pamela Raggi, Carmen Marciano, Rita Perna, Matteo Bassetti, Antonio Di Biagio, Maurizio Sanguinetti, Luca Masucci, Serafina Valente, Marco Mandalà, Alessia Giorli, Lorenzo Salerni, Patrizia Zucchi, Pierpaolo Parravicini, Elisabetta Menatti, Tullio Trotta, Ferdinando Giannattasio, Gabriella Coiro, Fabio Lena, Domenico A. Coviello, Cristina Mussini, Giancarlo Bosio, Enrico Martinelli, Sandro Mancarella, Luisa Tavecchia, Mary Ann Belli, Lia Crotti, Chiara Gabbi, Marco Rizzi, Franco Maggiolo, Diego Ripamonti, Tiziana Bachetti, Maria Teresa La Rovere, Simona Sarzi-Braga, Maurizio Bussotti, Stefano Ceri, Pietro Pinoli, Francesco Raimondi, Filippo Biscarini, Alessandra Stella, Mattia Bergomi, Kristina Zguro, Katia Capitani, Claudia Suardi, Simona Dei, Gianfranco Parati, Sabrina Ravaglia, Rosangela Artuso, Valentina Perticaroli, Antonio Perrella, Francesco Bianchi

The GEN-COVID is a multicentre academic observational study designed to collect and systematize biological samples and clinical data across multiple hospitals and healthcare facilities in Italy with the purpose of deriving patient-level phenotypic and genotypic data. The network started its activity on March 16, 2020 after IRB approval and since then more than 2000 patients have been collected and genotyped, in order to develop a genetic-based approach to understand the clinical variability of COVID-19 public health emergency. The data resulting from these studies are then stored and made available through the GEN-COVID Genetic Data Repository (GCGDR). The project collected and organized high-quality samples and data whose integrity was assured and could be readily accessed and processed for COVID-19 research using existing interoperability standards and tools. To this end, a GEN-COVID Biobank (GCB) and a GEN-COVID Patient Registry (GCPR) were established utilizing already existing biobanking and patient registry infrastructure.

**Clinical data**

Physicians of several specialties from Infectious Diseases to Anaesthesiology, Respiratory Diseases, Cardiology, Rheumatology, Neurology ENT specialty and Medical Genetics cooperated to create a detailed patient Registry and the biobank of samples. Patient data comprised demographic data, test results, diagnoses, and treatments were collected by the clinicians in charge of the patients and were stored in an anonymized excel file. The data resulting from GWAS study are stored and made available through the GEN-COVID Genetic Data Repository (GCGDR).

**Genotype data**

DNA extraction from peripheral blood (EDTA tube collection) after tween/SDS inactivation, using MagCore_HF16plus/ MagCore Genomic DNA Large Volume Whole blood kit. Quibit fluorometric quantification was performed. Genotyping and imputation were performed by the Genotyping laboratory of Institute for Molecular Medicine Finland FIMM Technology Centre, University of Helsinki. (Details are described in genotyping QC and imputation at FIMM section).

##### **Variability in immune response genes and severity of SARS-CoV-2 infection (≈)**

Anna M. Planas, Israel Fernández-Cadenas, Jordi Perez-Tur, Natalia Cullell, Laia Llucia-Carol, Alex Soriano, Veronica Rico, Daiana Aguero, Josep Lluis Bedini, Francisco Lozano, Carlos Domingo,Veronica Robles,Francisca Ruiz Jaén,Leonardo Márquez

The INMUNGEN-CoV2 study is a study of hospitalized SARS-CoV2 patients of age ranging from 18 to 65 years old, devoid of severe comorbidities and ongoing pathologies. This study was reviewed and approved by the Ethical Committee of the Hospital Clinic of Barcelona (CEIm number: Reg.HCB/2020/0357) and funded by the Spanish National Research Council (CSIC) (grant number 202020E086). We are indebted to the nursing staff at Hospital Clínic and to the patients for their generous donation, to Fundació Glòria Soler for its support to the COVIDBANK initiative and to the HCB-IDIBAPS Biobank for the biological human samples and data procurement.

**Clinical data**

Clinical data was obtained by accessing the individual clinical history of each patient through the Hospital centralized managing system by authorized personnel. We generated an excel file containing the needed information of each patient. The file was anonymized.

**Genotype data**

DNA was extracted from whole blood collected in EDTA tubes in a MagNA Pure LC 2.0 Instrument - Roche Life Science at the Centro de Diagnostico Biomedico of Hospital Clinic of Barcelona. Subsequent quality control (including removal of relatives or samples with low call rate) was performed in accordance with the data analysis plan provided by the COVID19-HGI^1^.

##### **Norwegian SARS-CoV-2 Study group (NorCoV2)**

Johannes R. Hov, Jan Cato Holter, Karl Erik Müller, Lars Heggelund, Andreas Lind, Fredrik Müller, Susanne Dudman, Aleksander Rygh Holten, Anne Margarita Dyrhol-Riise, Kristian Tonby, Jonas Bergan, Anders Benjamin Kildal, Vegard Skogen

Collection of SARS-CoV-2 PCR positive patients admitted to several Norwegian hospitals (Oslo University Hospital; Drammen Hospital, Hospital of North Norway Tromsø; Østfold Hospital Trust.), recruited by the Norwegian SARS-CoV-2 Study group. The project was reviewed and approved by the Regional Committee for Medical and Health Research Ethics in South-Eastern Norway (project no. 132550).

**Clinical data**

Clinical data were stored prospectively in study specific databases, and later retrieved for this study, complemented by retrospective addition of certain parameters.

**Genotype data**

DNA extraction from blood samples, genotyping and Imputation have been done at and by University of Kiel, Department of Molecular Medicine. (Details are described in genotype QC and imputation at Kiel section).

**Acknowledgements**

The following members of NorCov2 are also acknowledged: Synne Jenum, Oslo University Hospital; Børre Fevang, Oslo University Hospital; Birgitte Stiksrud, Oslo University Hospital; Else Quist-Paulsen, Oslo University Hospital; Simreen Kaur Johal, Oslo University Hospital; Anne Steffensen, Oslo University Hospital; Liv Hesstvedt, Oslo University Hospital; Dag Henrik Reikvam, Oslo University Hospital; Frank Pettersen, Oslo University Hospital; Vidar Ormaasen, Oslo University Hospital; Erik Egeland Christensen, Oslo University Hospital; Kjerstin Røstad, Oslo University Hospital; Linda Skeie, Oslo University Hospital; Marthe Jøntvedt Jørgensen, Oslo University Hospital; Sarah Nur, Oslo University Hospital; Gry Klouman Bekken, Drammen Hospital; Anne Hermann, Drammen Hospital; Hanne Opsand, Drammen Hospital; Bjørn Martin Woll, Drammen Hospital; Mette Bogen, Drammen Hospital; Cathrine Austad, Drammen Hospital; Garth Daryl Tylden, Hospital of North Norway Tromsø; Berit Gravrok, Hospital of North Norway Tromsø; Waleed Ghanima, Østfold Hospital Trust; Anne Marie Halstensen, Østfold Hospital Trust; Jorunn Brynildsen, Østfold Hospital Trust; Jeanette Aarem, Østfold Hospital Trust; Saad Aballi, Østfold Hospital Trust; Siri Øverstad, Østfold Hospital Trust; Kristine Marie Aarberg Lund, Østfold Hospital Trust; Åse-Berit Mathisen, Østfold Hospital Trust

##### **Determining the Molecular Pathways and Genetic Predisposition of the Acute Inflammatory Process Caused by SARS-CoV-2 (SPGRX)**

Marta E. Alarcón-Riquelme, Manuel Martínez-Bueno, David Bernardo, Silvia Rojo Rello

This is a study funded by the Andalusian Government (grant number CV20-10150). Collection was performed during March and April 2020. The protocol was reviewed and approved by the Valladolid Ethics Committee (PI-201716) and the Granada Ethics Committee (no number given) on March 24th, 2020 and April 13th, 2020, respectively.

**Clinical data**

Patients who arrived at the Valladolid hospital, Valladolid, Spain, with symptoms of covid-19 had a blood sample drawn, which was subsequently frozen. Patients were subjected to a PCR test and then hospitalized as deemed required. Thereafter clincal progression was monitored. Gender, age, and time of hospitalization, entry into ICU and/or death were reported.

**Genotype data**

DNA extraction was performed in Granada with standard procedures. Genotyping and imputation were performed at GENYO-Center for Genomics and Oncological Research. Genotyping was performed using the Illumina Global Screening Array-24 v3.0, and quality control (including removal of relatives or samples with low call rate) was performed in accordance with the data analysis plan provided by the COVID19-HGI^1^ (Specific parameters were described below).
Sample QC: call rate ≥ 0.98

Sample QC: Sex violations were excluded (those whose genetic sex does not match pedigree sex)

Sample QC: FHET (F coefficient for heterozygosity calculated by PLINK --het) within +/- 0.20

Sample QC: kinship coeficient threshold ≥ 0.2 (one sample of the pair excluded)

Sample QC: EUR ancestry (removed outliers defined by more than 8 SD in first two PCs)
Individuals with an average of more than 8 standard deviations of Euclidean distances calculated from the first two PCs

are considered outliers and are eliminated.

SNP QC: call rate ≥ 0.98

SNP QC: MAF ≥ 0.01

SNP QC: Hardy-Weinberg equilibrium (HWE) p value ≥ 1x10^-6^

The European ancestry of the samples was confirmed by comparison with the 1000 Genomes^3^ sub-populations by principal component analysis. Subsequent imputation was performed with Minimac4 on the Michigan Imputation Server using the European population of 1000 Genomes as reference panel. The minor allele frequency and imputation score for rs10490770 was 0.09 and 0.91, respectively.

##### **The genetic predisposition to severe COVID-19 (SweCovid)**

Hugo Zeberg, Robert Frithiof, Michael Hultström, Miklos Lipcsey and the Uppsala Intensive Care COVID-19 research group: Jacob, Rosén, Sarah Galien, Tomas Luther, Sara Bülow-Anderberg, Anna Gradin, Sten Rubertsson and Katja Hanslin

SweCovid is a study of PCR-verified COVID-19 patients admitted to an intensive care unit at Uppsala University Hospital, a university affiliated tertiary care center serving the population of the Uppsala-region in Sweden. The study was approved by the National Ethical Review Agency (EPM; 2020-01623). Written informed consent was obtained from patients, or next of kin if the patient was unable to give consent. The study was funded by the SciLifeLab/KAW national COVID-19 research program project grant to Michael Hultström (KAW 2020.0182), and the Swedish Research Council to Robert Frithiof (2014-02569 and 2014-07606). We are indebted to Elin Söderman, Joanna Wessbergh, Erik Danielsson and Philip Karlsson for excellent technical and administrative assistance.

**Clinical data**

Individual level patient data encompassing demographic data, test results, diagnoses, and treatments were obtained by medical chart review by trained clinicians or research coordinators. For biochemical and haematological tests, results were obtained by accessing the electronic medical record system database directly, rather than through chart reviews.

**Genotype data**

DNA from whole blood was extracted using the Indical Pathogen kit. 100 ul whole blood was diluted with 100 ul molecular grade water and DNA was extracted to a concentration of 15-90 ng/µl in a volume of 70 ul. DNA concentration was measured with Picodrop. The 70 ul was aliquoted and 40 ul was used for genotyping. Genotyping and imputation were performed by the Genotyping laboratory of Institute for Molecular Medicine Finland FIMM Technology Centre, University of Helsinki. (Details are described in genotyping QC and imputation at FIMM section).

##### **UK Biobank (UKB)**

Tomoko Nakanishi, Guillaume Butler-Laporte, Vincenzo Forgetta, J. Brent Richards

UK Biobank (UKB) is a population-based cohort which recruited people aged between 40 – 69 years old from across the UK, from 2006 to 2010. We first excluded subjects with no sex chromosome imputation results (N=291), those who withdrew consent recently (N=86), and those who died before March 16th, 2020. This resulted in a dataset of 457,941 individuals. UK Biobank study protocol and details in genotype QC are available online^7,8^. This study was conducted under project ID 27449. UK Biobank was approved by the Northwest Multi-Centre Research Ethics Committee and informed consent was obtained from all participants prior to participation.

We selected COVID-19 patients using the following strategy. We used the “covid19_result” table to obtain SARS-CoV-2 PCR test results and inpatient hospital data (“hesin” tables; admitted patient care in England, available up to Dec 9^th^, 2020) to obtain COVID-19 diagnosis data and associated phenotypes, as well as the death register data (“death” and “death_cause” tables) to obtain mortality information (available up to Dec 17^th^, 2020). We restricted the analysis to the period ranging from March 16th, 2020 to Dec 17^th^, 2020. A diagnosis of COVID-19 was done according to 1) at least one SARS-CoV-2 PCR positive test results from “covid19_result” table, and/or 2) ICD-10 codes of “U071” (COVID-19 virus identified) or “U072” (COVID-19 virus not identified) from “hesin” tables. (N=8,328) All codes to generate this data are available on [github](https://github.com/tomoconaka/COVID19-chr3).

**Phenotype definitions**

All codes to generate this data are available on [github](https://github.com/tomoconaka/COVID19-chr3).

**Demographics**

Age at COVID-19 diagnosis was obtained by using the age when attended assessment centre (data-field: 21003), date of attending assessment centre (data-field: 53) and the date of COVID-19 diagnosis. The dates of COVID-19 diagnosis were obtained either from “covid19_result” table or from the hospitalization date from “hesin” table if missing.

We used data-field: 31 for sex, height (data-field: 50) and weight (data-field: 21002) for the calculation of the BMI. Smoking status was defined by the questionnaire - based information as was previously reported^9^. “Never smokers” were people who answered “not smoking at present” and “never smoked in the past”, or who answered “not smoking at present, smoked occasionally or just tried once or twice in the past” but didn’t have more than 100 episodes of smoking over their lifetime. “Current smokers” were those who smoke at present on most, or all days, or occasionally. “Past smokers” were those who do not smoke at present but smoked on most or all days in the past, who do not smoke at present and smoked occasionally or just tried once or twice in the past but had at least 100 episodes of smoking over their lifetime, or who smoked on most or all days or occasionally with more than 100 episodes of smoking over their lifetime but prefer not to answer whether they smoke at present. (Data-fields 1239, 1249 and 2644 were used)

**Comorbidities**

We defined the following comorbidities using data-tables available in UKB; transplantation, diabetes mellitus, chronic obstructive pulmonary disease, chronic kidney disease, chronic heart failure, and cancer. Codes used for disease ascertainment are listed in **Supplementary Table 15**.

**Hospital complications**

Hospitalization was defined by the following criteria. 1) SARS-CoV-2 PCR positive individuals with “hospital inpatient” flag (reqorg = 1 in UKB) when they were tested 2)

patients with at least one inpatient hospital episode within 30 days after COVID-19 diagnosis and 3) patients with ICD-10 codes of “U071” or “U072”.

ICU admission was defined by at least one critical care episode within 30 days after COVID-19 diagnosis using “hesin_critical” table and ICU duration was calculated using the starting and ending dates of the critical care episode.

Hospital complications below refer to events that were diagnosed within 30 days after COVID-19 diagnosis or the events that were recorded in the same episodes with ICD-10 codes of “U071” or “U072”. These events were treated as missing in patients diagnosed with COVID-19 after Dec 9^th^ since inpatient hospital data (“hesin” tables; admitted patient care in England) was available only up to Dec 9^th^, 2020.

Severe respiratory failure was defined as 1) being on a ventilator at least one day according to “hesin_critical” table (aressupdays > 0) 2) with ICD-10 codes of adult respiratory distress syndrome (ARDS) ("J80"), acute respiratory failure ("J9600","J9609"), dependence on ventilator ("Z991") or OPCS codes of the use of ventilator ("E851","E852"). Days on a ventilator were also obtained by “aressupdays”. Respiratory failure was defined by including being on oxygen therapy in ICU (bressupdays > 0 in “hesin_critical” table) in addition to severe respiratory failure above. Venous thromboembolism was defined by having ICD-10 codes of venous embolism and thrombosis (“I81”, “I82*”), or Pulmonary embolism (I26*). Cardiovascular complications was defined by having ICD-10 codes of stroke (“I61”, “I62”, “I63”, “I64”, “I65”, ”I66*”) or myocardial infarction (“I21*”). Renal complications was defined by having ICD-10 codes of acute kidney injury (“N17*”). Hepatic complications was defined by having ICD-10 codes of acute hepatic failure (“K720”).

**Death information**

For mortality analysis, those who were deceased with ICD-10 codes of “U071” and/or “U072” as causes of death were treated as COVID-19 related death. Otherwise, we treated them as unrelated death. Those without any death register were assumed to be alive until Dec 17^th^, 2020.

#### **Genotype QC and imputation at FIMM**

Mattia Cordioli, Mari Niemi, Lindo Nkambul, Sara Pigazzini

The Genotyping Laboratory of Institute for Molecular Medicine Finland, University of Helsinki provided genotyping for the following cohorts: BeLCovid_1, BeLCovid_2, GEN-COVID and SweCovid. Genotyping was performed with Illumina Global Screening Array-24 v3.0 + Multi-Disease beadchip.

Quality control was performed for the above-mentioned studies and for the BRACOVID data, genotyped with Axiom_PMRA.r3 array and thus analyzed separately. Quality control was performed in accordance with the data analysis plan provided by the COVID19-HGI^1^ and as described below.

Sample QC: samples were removed with the following criteria:

- call rate ≤ 0.98
- duplicated samples (IBD > 0.98)
- genetically inferred sex not matching the reported sex and could not be confirmed by the collection center

SNP QC: SNPs were removed with the following criteria:

- call rate ≤ 0.98
- biallelic and strand-ambiguous SNPs
- Hardy-Weinberg equilibrium (HWE) p value ≥1x10^-10^. HWE was tested separately in each cohort. HWE for chromosome X was tested considering only females.
- allele frequency (AF) deviating from the Genome Aggregation Database (gnomAD)^10^ ancestry-specific reference panel. Difference was calculated using Mahalanobis distance (MD) and excluding SNPs with MD>30.

Genotype imputation was conducted for chromosomes 1-22 and X data using the novel TOPMed Freeze5^4,5^ on genome build GRCh38 and the Michigan Imputation Server^6^.

#### **Genotype QC and imputation at Kiel (COVID19-Host(a)ge)**

David Ellinghaus, Frauke Degenhardt, Luis Bujanda, Maria Buti, Agustín Albillos, Pietro Invernizzi, Javier Fernández, Daniele Prati, Guido Baselli, Rosanna Asselta, Marit M. Grimsrud, Chiara Milani, Fátima Aziz, Jan Kässens, Sandra May, Mareike Wendorff, Lars Wienbrandt, Florian Uellendahl-Werth, Tenghao Zheng, Xiaoli Yi, Raúl de Pablo, Adolfo G. Chercoles, Adriana Palom, Alba-Estela Garcia-Fernandez, Francisco Rodriguez-Frias, Alberto Zanella, Alessandra Bandera, Alessandro Protti, Alessio Aghemo, Ana Lleo, Andrea Biondi, Andrea Caballero-Garralda, Andrea Gori, Anja Tanck, Anna Carreras Nolla, Anna Latiano, Anna Ludovica Fracanzani, Anna Peschuck, Antonio Julià, Antonio Pesenti, Antonio Voza, David Jiménez, Beatriz Mateos, Beatriz Nafria Jimenez, Carmen Quereda, Cinzia Paccapelo, Christoph Gassner, Claudio Angelini, Cristina Cea, Aurora Solier, David Pestaña, Eduardo Muñiz-Diaz, Elena Sandoval, Elvezia M. Paraboschi, Enrique Navas, Félix García Sánchez, Ferruccio Ceriotti, Filippo Martinelli-Boneschi, Flora Peyvandi, Francesco Blasi, Luis Téllez, Albert Blanco-Grau, Georg Hemmrich-Stanisak, Giacomo Grasselli, Giorgio Costantino, Giulia Cardamone, Giuseppe Foti, Serena Aneli, Hayato Kurihara, Hesham ElAbd, Ilaria My, Iván Galván-Femenia, Javier Martín, Jeanette Erdmann, Jose Ferrusquía-Acosta. Koldo Garcia-Etxebarria, Laura Izquierdo-Sanchez, Laura R. Bettini, Lauro Sumoy, Leonardo Terranova, Leticia Moreira, Luigi Santoro, Luigia Scudeller, Francisco Mesonero, Luisa Roade, Malte C. Rühlemann, Marco Schaefer, Maria Carrabba, Mar Riveiro-Barciela, Maria E. Figuera Basso, Maria G. Valsecchi, María Hernandez-Tejero, Marialbert Acosta-Herrera, Mariella D’Angiò, Marina Baldini, Marina Cazzaniga, Martin Schulzky, Maurizio Cecconi, Michael Wittig, Michele Ciccarelli, Miguel Rodríguez-Gandía, Monica Bocciolone, Monica Miozzo, Nicola Montano, Nicole Braun, Emanuele Pontali, Nilda Martínez, Onur Özer, Orazio Palmieri, Paola Faverio, Paoletta Preatoni, Paolo Bonfanti, Paolo Omodei, Paolo Tentorio, Pedro Castro, Pedro M. Rodrigues, Aaron Blandino Ortiz, Rafael de Cid, Ricard Ferrer, Roberta Gualtierotti, Rosa Nieto, Siegfried Goerg, Salvatore Badalamenti, Sara Marsal, Giuseppe Matullo, Serena Pelusi, Simonas Juzenas, Stefano Aliberti, Valter Monzani, Victor Moreno, Tanja Wesse, Tobias L. Lenz, Tomas Pumarola, Valeria Rimoldi, Silvano Bosari, Wolfgang Albrecht, Wolfgang Peter, Manuel Romero-Gómez, Mauro D’Amato, Stefano Duga, Jesus M. Banales, Johannes R Hov, Trine Folseraas, Luca Valenti, Andre Franke, Tom H. Karlsen

Data released for this project are based on Spanish (COVID-19 Hostage_1, COVID-19 Hostage_2, COVID-19 Hostage_3), Italian (COVID-19 Hostage_4, FoGS) and Norwegian (NorCoV2) case-controls GWAS datasets used for genome-wide discovery analysis of SNPs associated with severe-COVID19. Release 1 of this project was recently published under the consortium denomination of “Severe Covid-19 GWAS group”^11^, yet is identical to Host(a)ge in terms of Covid-19 Host Genetics Consortium participation.

**Genotyping & Genotype calling**

Genotyping of all patient panels and the control panels was conducted at the Institute of Clinical Molecular Biology’s DNA Laboratory and Genotyping Core Facilities, employing Illumina’s (Illumina Inc., San Diego, U.S.) Global Screening Array-24 Multi Disease (GSA) Version 2.0 B1 following the Illumina(R) Infinium HTS Assay Auto 3-day Workflow (Document #15045738v0). Initial genotype calling extracting GSA genotyped data from intensity data files was performed with the Illumina GenomeStudio v. 2.0 software with the cluster definition files [GSAMD24v2-0_20024620_A1-762Samples-LifeBrain](https://www.illumina.com/documents/products/datasheets/datasheet_genomestudio_software.pdf). Finally, we had 712,189 SNPs before quality control (QC). Based on initial genotype data, we removed samples with <90% call rate using PLINK^12^.

**Genotype QC**

After genotype calling a unified QC procedure was carried out for the Spanish, Italian and Norwegian case-controls GWAS datasets. Variants that had >2% missing data, a MAF <0.1% in disease sets or in controls, different missing genotype rates in affected and unaffected individuals (*P_Fisher_*<10^-5^) or deviated from HWE (with a false discovery rate (FDR) threshold of 10^-5^ in controls) (a) across the entire collection with at most one batch being removed or (b) falling below in two single batches, were excluded. Samples that had overall increased/decreased heterozygosity rates (i.e. ±5 SD away from the sample mean) were removed. For robust duplicate/relatedness testing (IBS/IBD estimation) and population structure analysis, we used a linkage disequilibrium (LD)-pruned subset of SNPs on the basis of a set of independent (MAF>5%) SNPs excluding X- and Y-chromosomes, SNPs in LD (leaving no pairs with r^2^>0.2), and 11 high-LD regions as described by Price *et al*^13^*.* Pairwise percentage IBD values were computed using PLINK^12^.

One individual (the one showing greater missingness) from each pair with PI_HAT>0.1875 was removed. To identify ancestry outliers, i.e. subjects of non-European ancestry, to resolve within-Europe relationships and to test for population stratification within and across batches (merged for the Italian, Spanish and Norwegian panels), we performed principal component analysis (PCA) for remaining QCed cases and controls using the PCA method, as implemented in FlashPCA^14^, using a LD-pruned subset of SNPs (see text above). The QCed Italian, Spanish and Norwegian GWAS datasets comprised contained 525,788 (Italy), 522,416 (Spain), and 525,836 (Norway) variants after QC and filtering of SNPs with alleles AT or CG (the latter often leading to strand issues during imputation).

**Extraction of non-European individuals**

Ancestry outliers not matching European populations during QC (see above) were extracted from the pre-QC genotype data. Variants present in all post-QC populations were extracted from the data. We determined relatedness as described above and removed one (the one showing greater missingness) from each pair with PI_HAT>0.1875.

**SNP genotype imputation**

Genotype imputation was conducted for chromosomes 1-22 and X data using the novel TOPMed Freeze5^5^ on genome build GRCh38 and the [Michigan Imputation Server](https://imputation.biodatacatalyst.nhlbi.nih.gov/index.html)^6^. We provided the input data in “vcf.gz” format as GRCh38 build (European dataset) and GRCh37 (non- European dataset). We used the offered population panel “ALL” (European dataset) and skipped QC-checks for the non-European dataset. We applied the server-side option to filter by an imputation R^2^ with threshold 0.1. We then extracted the chr3 variant rs10490770 which had a post-imputation R^2^ >0.92 in all populations.

#### **Individual study information of external cohorts**

##### **FinnGen**

Sara Pigazzini, Mattia Cordioli, Mari Niemi

FinGenn is a public-private partnership project combining genotype data from Finnish biobanks and digital health record data from Finnish health registries (https://www.finngen.fi/en). Six regional and three country-wide Finnish biobanks participate in FinnGen. FinnGen also includes data from previously established populations and disease-based cohorts. Ethical approval was obtained by HUS coordinating Ethics Committee.

**Clinical data**

The data release contains information on coronavirus positive individuals in the Finnish National Infectious Diseases Register.

Data is updated from the register monthly at the moment. The data of this version (R6_7.0) has been collected from the register on 01.02.2021. Other phenotypic data used as covariates in the study were obtained from Finnish national and health care registers.

**Genotype data**

Samples were genotyped with Illumina (Illumina Inc., San Diego, CA, USA) and Affymetrix arrays (Thermo Fisher Scientific, Santa Clara, CA, USA). Genotype calls were made with GenCall and zCall algorithms for Illumina and AxiomGT1 algorithm for Affymetrix data. Chip genotyping data produced with previous chip platforms and reference genome builds were lifted over to build version 38 (GRCh38/hg38) following the protocol described elsewhere^15^. In sample-wise quality control, individuals with ambiguous sex, high genotype missingness (>5%), excess heterozygosity (+-4SD) and non-Finnish ancestry were removed. In variant-wise quality control variants with high missingness (>2%), low HWE P-value (<1e-6) and minor allele count, MAC<3 were removed. Chip genotyped samples were pre-phased with Eagle 2.3.5 (https://data.broadinstitute.org/alkesgroup/Eagle/) with the default parameters, except the number of conditioning haplotypes was set to 20,000. Genotype imputation was done with the population-specific SISu v3 reference panel. High-coverage (25-30x) WGS data (N= 3,775) were generated at the Broad Institute and at the McDonnell Genome Institute at Washington University; and jointly processed at the Broad Institute. Variant call set was produced with GATK HaplotypeCaller algorithm by following GATK best-practices for variant calling. Genotype-, sample- and variant-wise QC was applied in an iterative manner by using the Hail framework (https://github.com/hail-is/hail) v0.1 and the resulting high-quality WGS data for 3,775 individuals were phased with Eagle 2.3.5 as described above. Genotype imputation was carried out by using the population-specific SISu v3 imputation reference panel with Beagle 4.1 (version 08Jun17.d8b, https://faculty.washington.edu/browning/beagle/b4_1.html) as described elsewhere^16^. Post-imputation quality-control involved non-reference concordance analyses, checking expected conformity of the imputation INFO-values distribution, MAF differences between the target dataset and the imputation reference panel and checking chromosomal continuity of the imputed genotype calls.

**Analysis details**

Three COVID-19 severity outcomes (hospitalization, ICU admission and death or severe respiratory failure) and one COVID-19 complications (severe respiratory failure) were available at FinnGen. We performed logistic regressions for these four outcomes on rs10490770 risk allele carrier status adjusted for age, sex, age^2^, an age by sex interaction term and first 10 PCs. We then also performed linear regressions between age and rs10490770 risk allele carrier status adjusted for sex and first 10 PCs amongst cases for three COVID-19 severity outcomes.

##### **Columbia University COVID-19 Biobank (CUB)**

Atlas Khan, Ning Shang, Chen Wang, Gundula Povysil, Nitin Bhardwaj, Sheila M. O’Byrne, Renu Nandakumar, Amritha Menon, Yat S. So, Danielle Pendrick, Eldad Hod, Ali G. Gharavi, luliana Ionita-Laza, Soumitra Sengupta, Wendy Chung, Muredach P. Reilly, David B. Goldstein, Krzysztof Kiryluk

The Columbia University COVID-19 Biobank was established in response to the New York City infection surge in March 2020. The biobank recruited COVID-19 cases of diverse ancestry among all patients who were treated at Columbia University Irving Medical Center between March and May 2020. All cases were diagnosed by positive SARS-CoV-2 PCR test based on nasopharyngeal samples. Ethical approval was obtained from the Columbia University IRB.

**Clinical data**

Individual level patient data encompassing demographic data, test results, diagnoses, and treatments were obtained by medical chart review by trained clinicians or research coordinators. The mean age of COVID-19 patients was 62.89 years, and the percentage of females was 43%. Overall, the cohort was composed of 508 (49%) patients of European, 332 (32%) of African American, and 189 (18%) of Hispanic/Latinx ancestry. Because of hospital-based ascertainment, the cohort was enriched in more severe disease with 938 (91%) patients being hospitalized for COVID-19 or its complications, 267 (26%) patients receiving non-invasive ventilation for COVID-19 pneumonia, and 201 (25%) patients requiring intubation with mechanical ventilation for COVID-19 respiratory failure. The overall mortality rate from COVID-19 was 24% in this cohort.

**Genotype data**

DNA of whole blood samples was extracted using standard procedures and genotyping was performed using the Illumina Global Diversity Array (GDA) chip at the Columbia University Institute for Genomic Medicine (IGM). The analysis of intensity clusters and genotype calls were performed in Illumina Genome Studio software; all SNPs were called on forward DNA strand and standard QC filters were applied, including per-SNP genotyping rate >95%, per-individual genotyping rate >90% and MAF>0.01. The duplicates and cryptic relatedness in the given cohort were determined and excluded based on the estimated pairwise kinship coefficients >0.0884. After quality control analyses, 1,029 cases genotyped for 1,096,321 SNPs with overall genotyping rate of 99.9% were included in downstream analyses. Genome-wide imputation was performed using the TopMed imputation server^6^. A total of 13,439,413 common (MAF>0.01) markers were imputed at high quality (R2>0.8) and included in the analysis.

**Analysis details**

Cases with death or severe respiratory failure phenotype were defined as those either deceased, requiring non-invasive ventilation, or requiring intubation with mechanical ventilation. Cases with severe respiratory failure phenotype were defined as those requiring non-invasive ventilation, or requiring intubation with mechanical ventilation regardless of their vital status. We performed linear regression between age at diagnosis and the carrier status adjusted for sex and first five principal components of ancestry. The analyses were performed within each genetically defined major continental ancestry group.

### Supplementary methods and results

#### **Population assignment and genetic principal components calculations**

To infer continent-wise ancestries of individuals in COVID-19 HGI, we first combined individual-level genotype data available from all 17 cohorts available and filtered HapMap3^17^ SNPs with MAF>0.1 which were shared with 1000 Genomes (1000G) phase3^3^ marker panel. We also excluded variants within high LD regions (“chr6:25-35Mb” and “chr8:7-13Mb” on GRCh38). This resulted in 71,007 variants shared between all panels. We first performed PCA using a reference panel (1000G) and subsequently projected all samples onto the 1000G PCA space. We next performed a random forest classifier-based approach by training on a reference panel (five different super populations of 1000G; EUR, AFR, SAS, EAS and AMR, in total 2,003 samples [80% of samples]) for training the random forest models with ntree=100, mtry=sqrt(71,007), min nodesize=2 using RandomForestClassifier from sklearn in python and assigning ancestries to the individuals of interest in our study. (**Supplementary Figure 1**) We used a relaxed threshold of 0.5 as a cut-off of the probability of being in each population. Thus, some individuals in each ancestry could have admixed components of ancestries. All of this analysis was done using “hail v0.2.61”^18^ and the codes were described in [github](https://github.com/tomoconaka/COVID19-chr3). The first five PCs were used as covariates and were standardized when convergence was failed in each regression model.

###### Supplementary Figure 1: Bivariate scatterplots of the genetic principal components

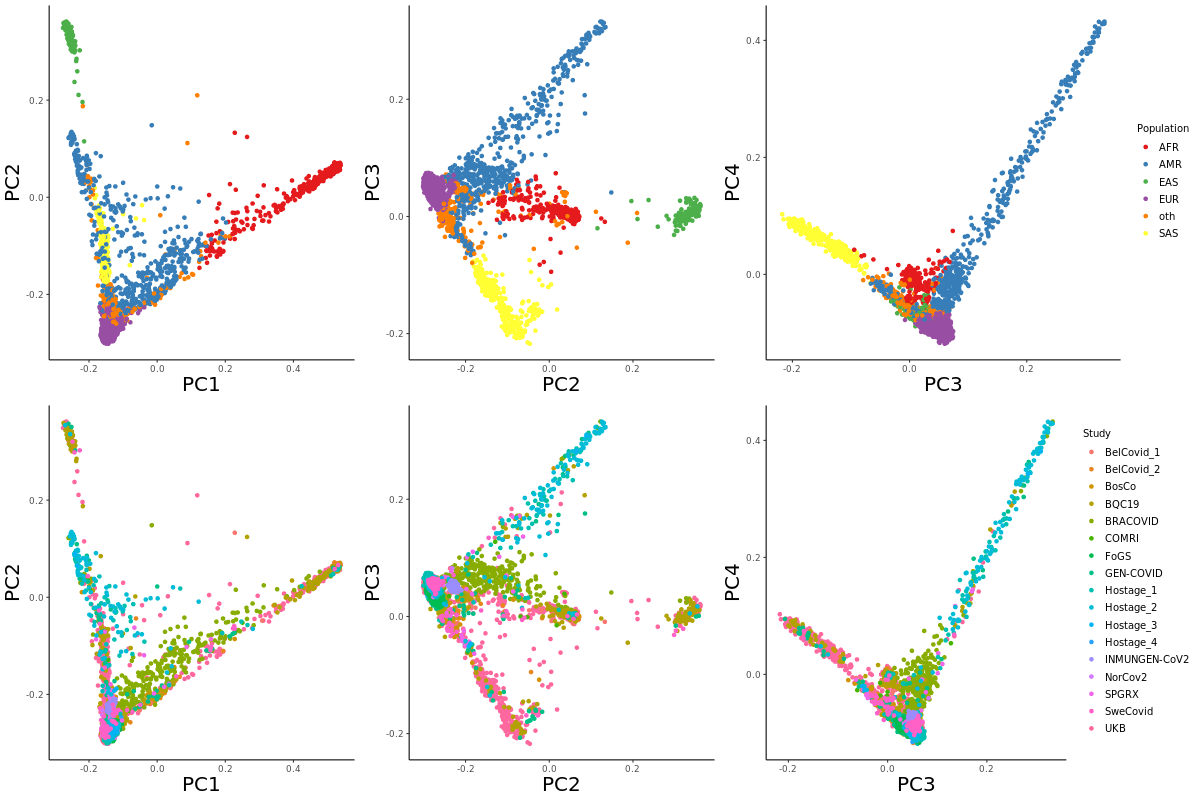

#### **Genetic model selection**

For genetic model selection, we compared three models (dominant, recessive and additive)^19^ using mixed effects logistic regression analysis for death or severe respiratory failure using “lme4 v1.1-25” R package and the codes were described at [github](https://github.com/tomoconaka/COVID19-chr3). These analyses were conducted specifically in individuals of European descent. All analyses were conducted on the rs10490770 risk allele carrier status and adjusted for age, sex, and the first five genetic PCs, calculated as described above, as fixed effects and groups indicating participating studies as random effects. **Supplementary Table 2** summarizes the overall comparison of the models by the Akaike Information Criteria (AIC), the Bayesian information criteria (BIC), log-likelihood of the models and deviance of the residuals. We chose to apply a dominant model since this model estimated to have the lowest error of prediction.

#### **Defining the study groups to account for the study specific variability**

As discussed in the main text, each cohort had its own strategy to recruit patients and ascertain severity of COVID-19 and COVID-19 complications. Since this may lead to bias our estimates, we decided to adjust for the participating studies as random effects. However, some cohorts were enriched for severe cases intentionally, and including these studies as covariates may induce collinearity and affect statistical power. To alleviate these issues, we first grouped the participating studies from the same country into one group; Belgium, Brazil, Canada, Germany, Italy, Norway, Spain, Sweden, and UK. Since this step would have left some groups with no controls for death or severe respiratory failure phenotypes, we additionally combined Brazil, Canada, Germany, Norway and Sweden to the same group, resulting in five different groups to be included as covariates assuming random effects. This strategy may not fully adjust for the study specific variability, and we therefore also included the participating studies themselves as covariates in the sensitivity analyses.

#### **Association with laboratory values**

**Data quality control and outlier removal**

We first collected longitudinal laboratory values which were known to be associated with severe COVID-19^20–24^ and harmonized the units of these values across all cohorts. The final list of tested laboratory values included white blood cell counts, lymphocyte counts, neutrophil counts, monocyte counts, platelet counts, C-reactive protein, Troponin T, aspartate aminotransferase, alanine aminotransferase, total bilirubin, lactate dehydrogenase, γ-glutamyl transpeptidase, alkaline phosphatase, D-dimer, interleukin 6, ferritin, procalcitonin, and creatine kinase, fibrinogen and creatinine. Some laboratory values were excluded from the analysis. For example, CD4 and CD8 counts had too small sample sizes to analyze. Eosinophil and basophil counts were too low to be detected in some cohorts’ biochemistry systems, leading to many “zero” values, and a lack of consistency. Values which exceeded the higher measurable limit or which were below the lower measurable limit were set to the higher or lower limit values respectively.

To account for local measurement effects, we first imputed zero values to the lowest measurement amongst all individuals divided by two. We then natural-log transformed the values and regressed out the participating studies as fixed effects using linear regression models. We applied a median-based outlier detection method by considering an observation as being an outlier if the absolute difference between the observation and the sample median is larger than the five times median absolute deviation divided by 0.6745 (0.75^th^ quantile of the standard normal distribution) (**Supplementary Figure 2**). The codes were described at [github](https://github.com/tomoconaka/COVID19-chr3).

###### Supplementary Figure 2: Laboratory values of pre- and post-QC.

The top figures: (x-axis) individual id, (y-axis) natural-log transformed values adjusted for the participating studies.
The bottom figures: (x-axis) study id (y-axis) post-QC raw laboratory values.

lab_wbc: white blood cell counts, lab_lymphocytes: lymphocyte counts, lab_neutrophils: neutrophil counts, lab_ monocytes: monocyte counts, lab_platelets: platelet counts, lab_crp: C-reactive protein, lab_trop_t: Troponin T, lab_trop_i: Troponin I, lab_ast: aspartate aminotransferase, lab_alt: alanine aminotransferase, lab_bilirubin: total bilirubin, lab_ldh: lactate dehydrogenase, lab_ggt: γ-glutamyl transpeptidase, lab_alp: alkaline phosphatase, lab_d_dimer: D-dimer, lab_il_6: interleukin 6, lab_serum_ferritin: Ferritin, lab_procalcitonin: procalcitonin, lab_ck: creatine kinase, lab_fibrinogen: Fibrinogen, lab_creatinine: creatinine

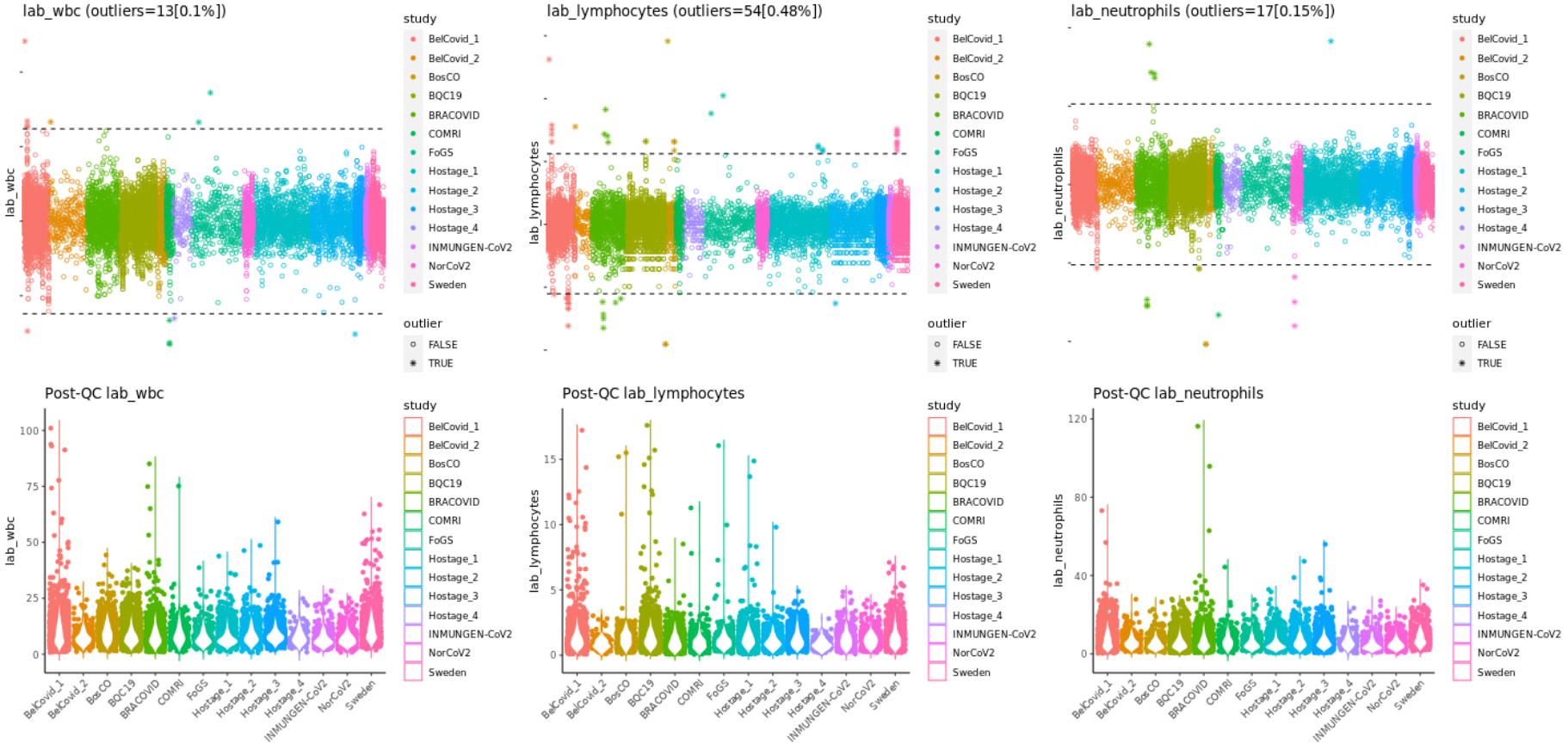

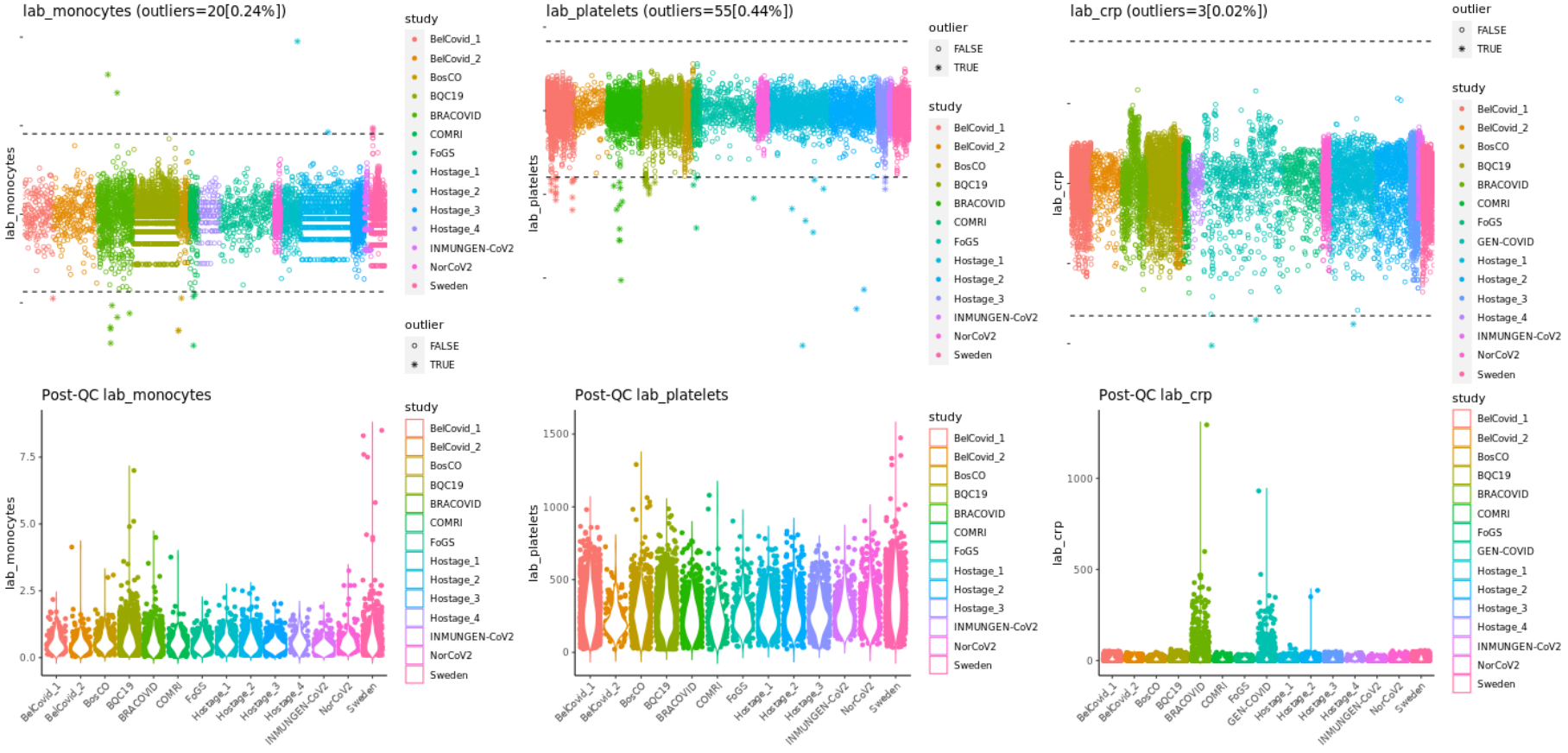

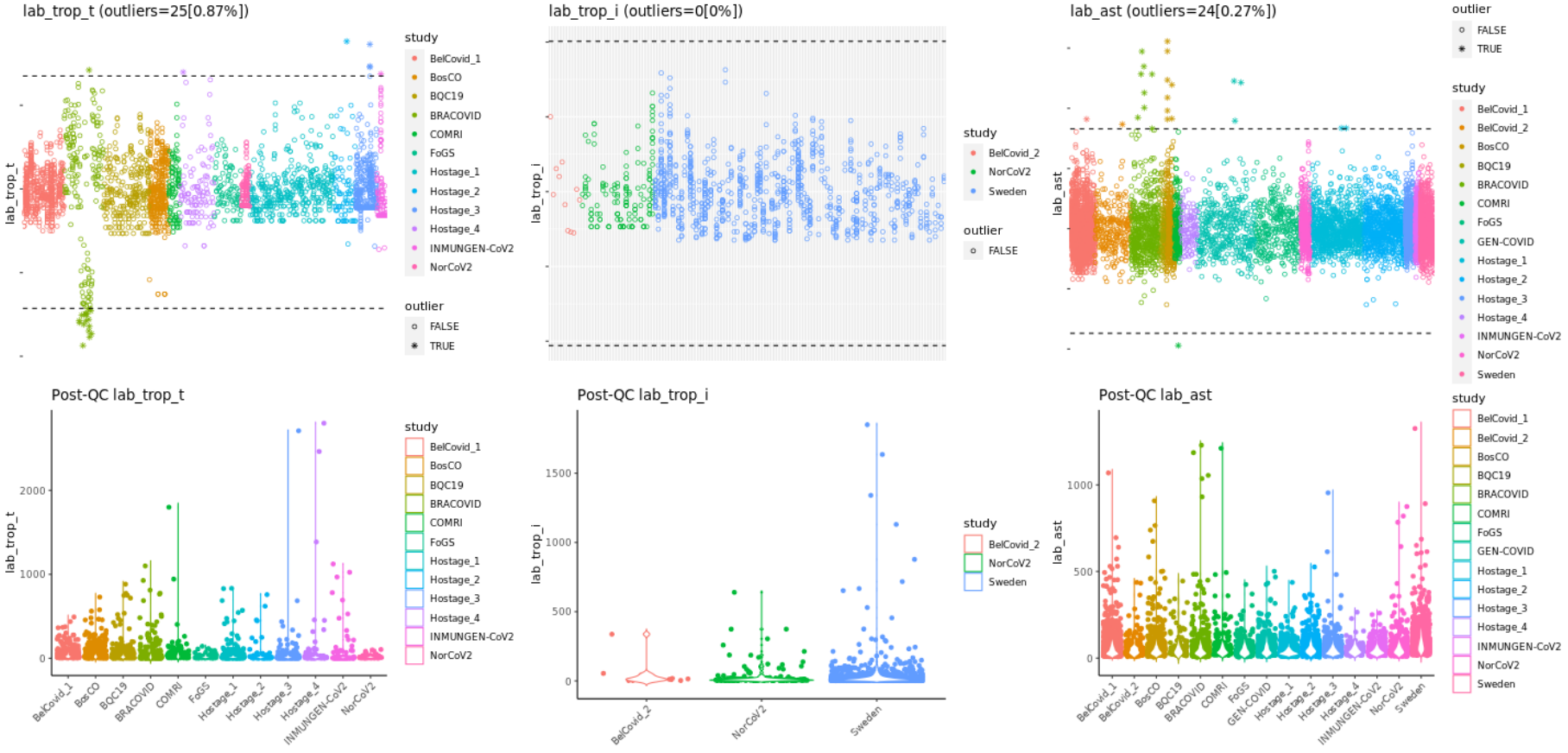

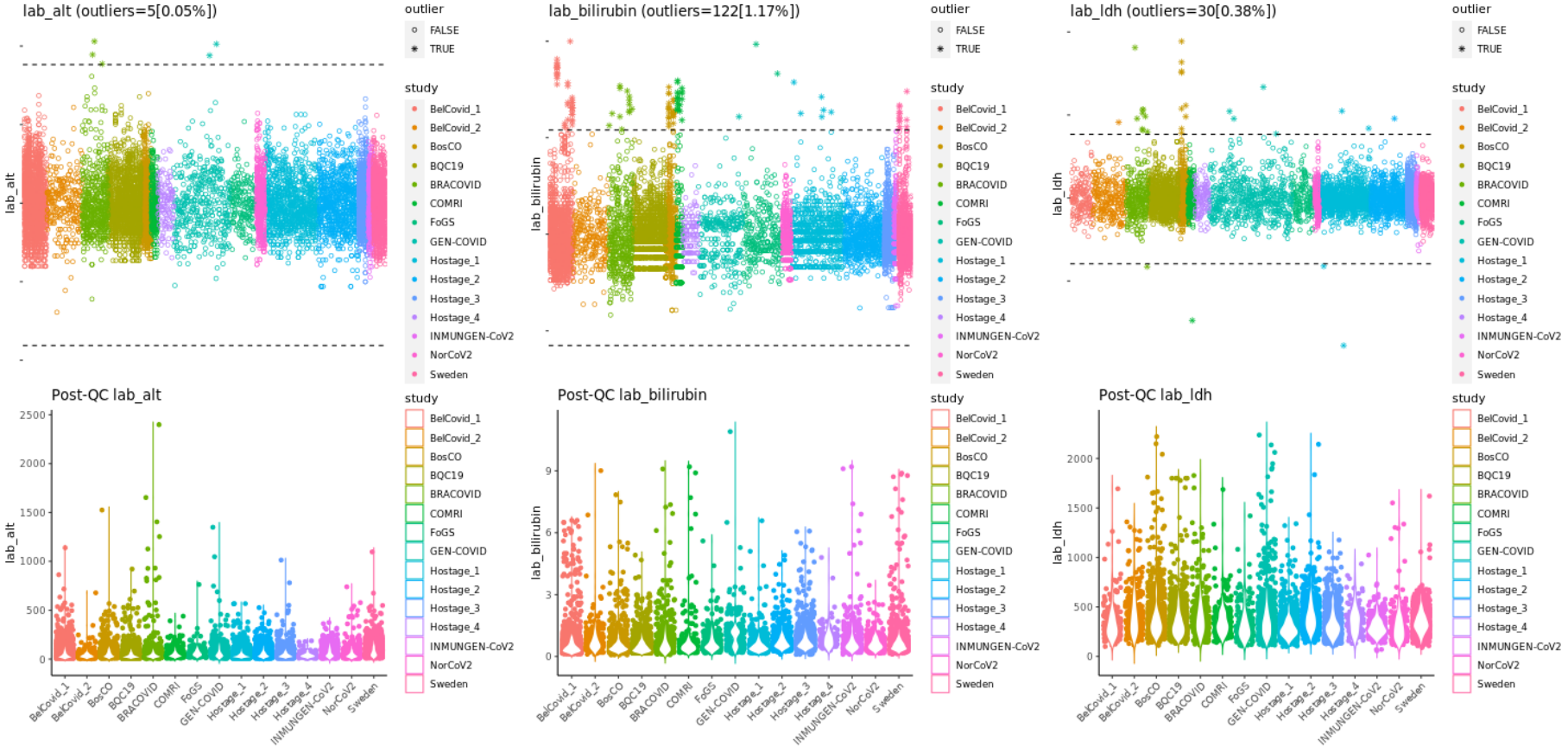

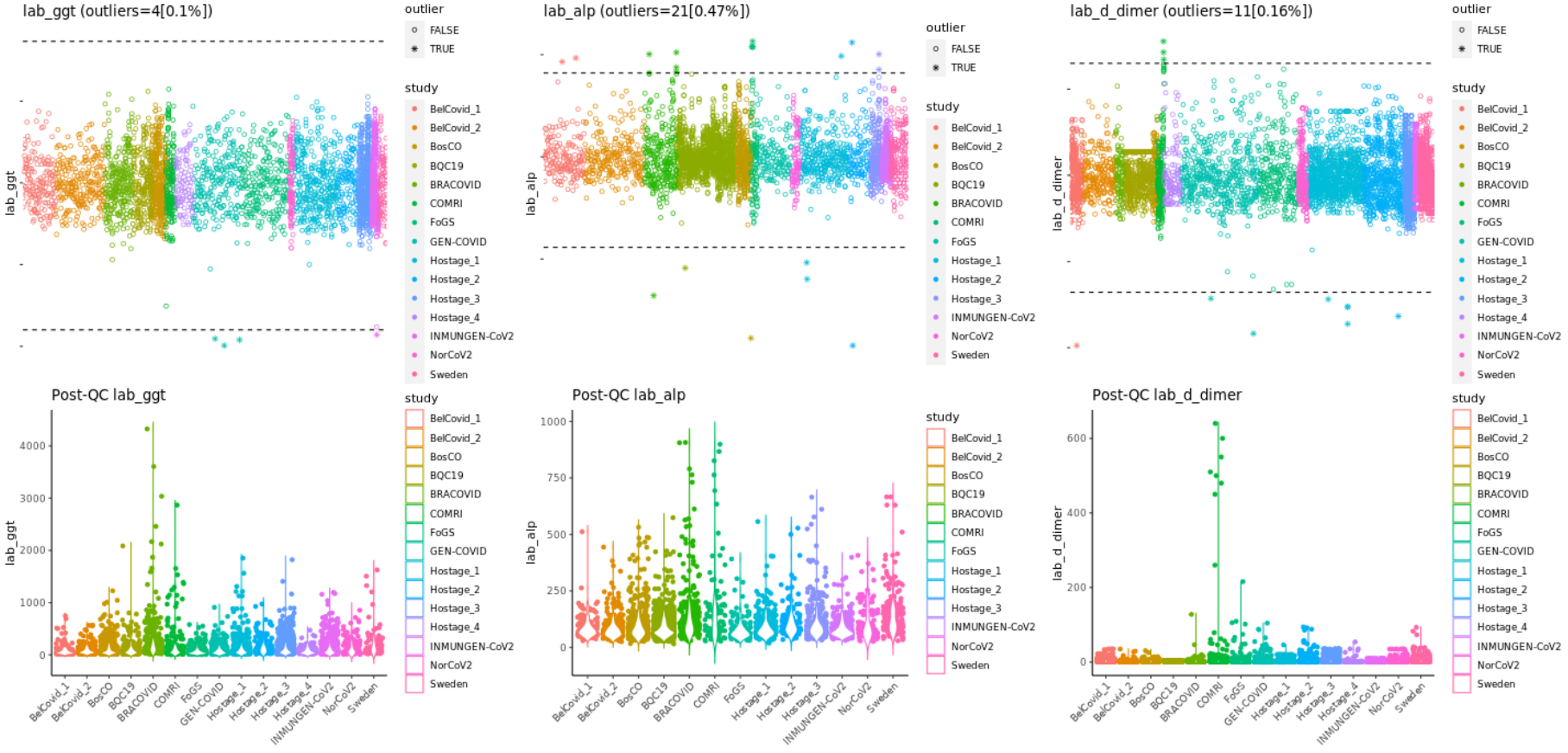

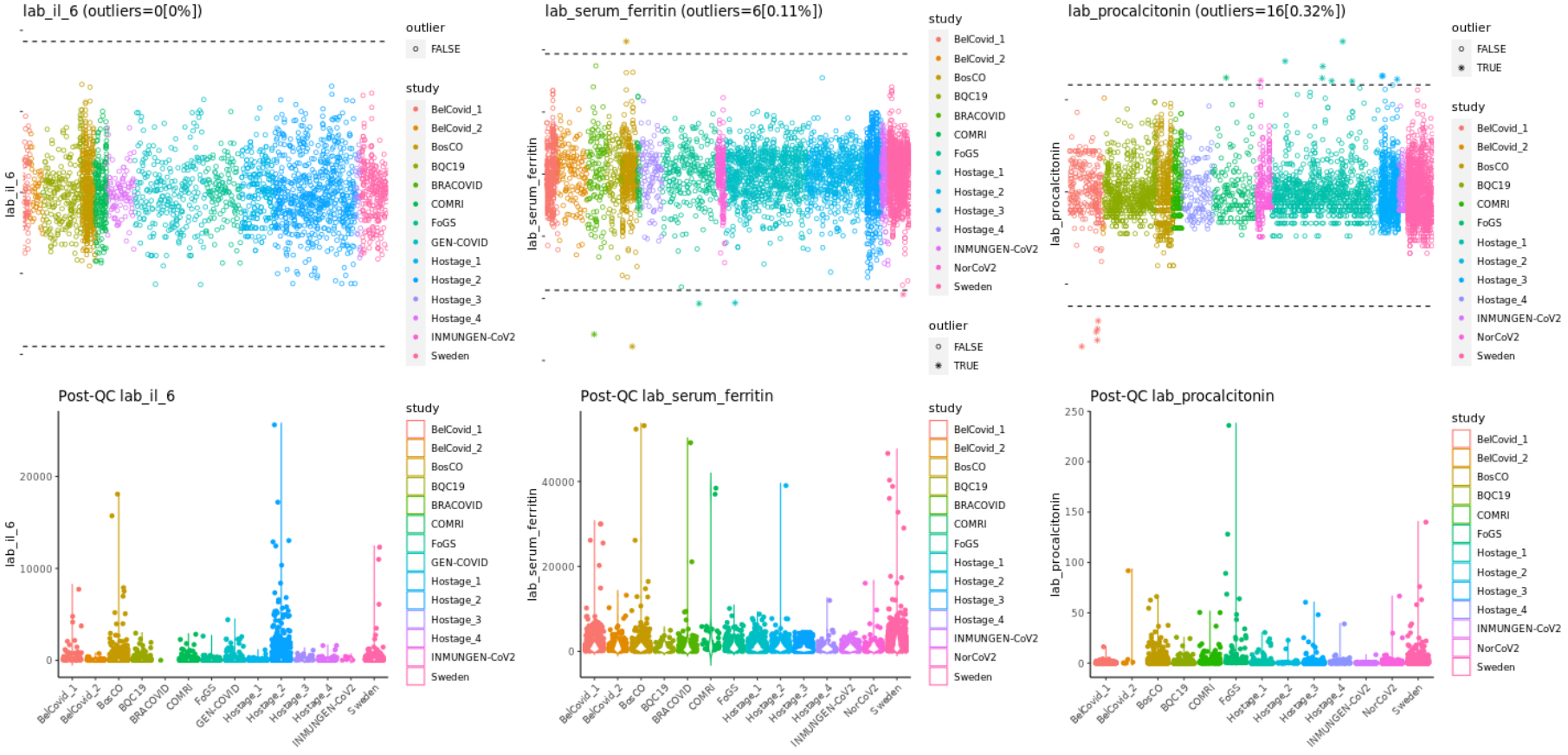

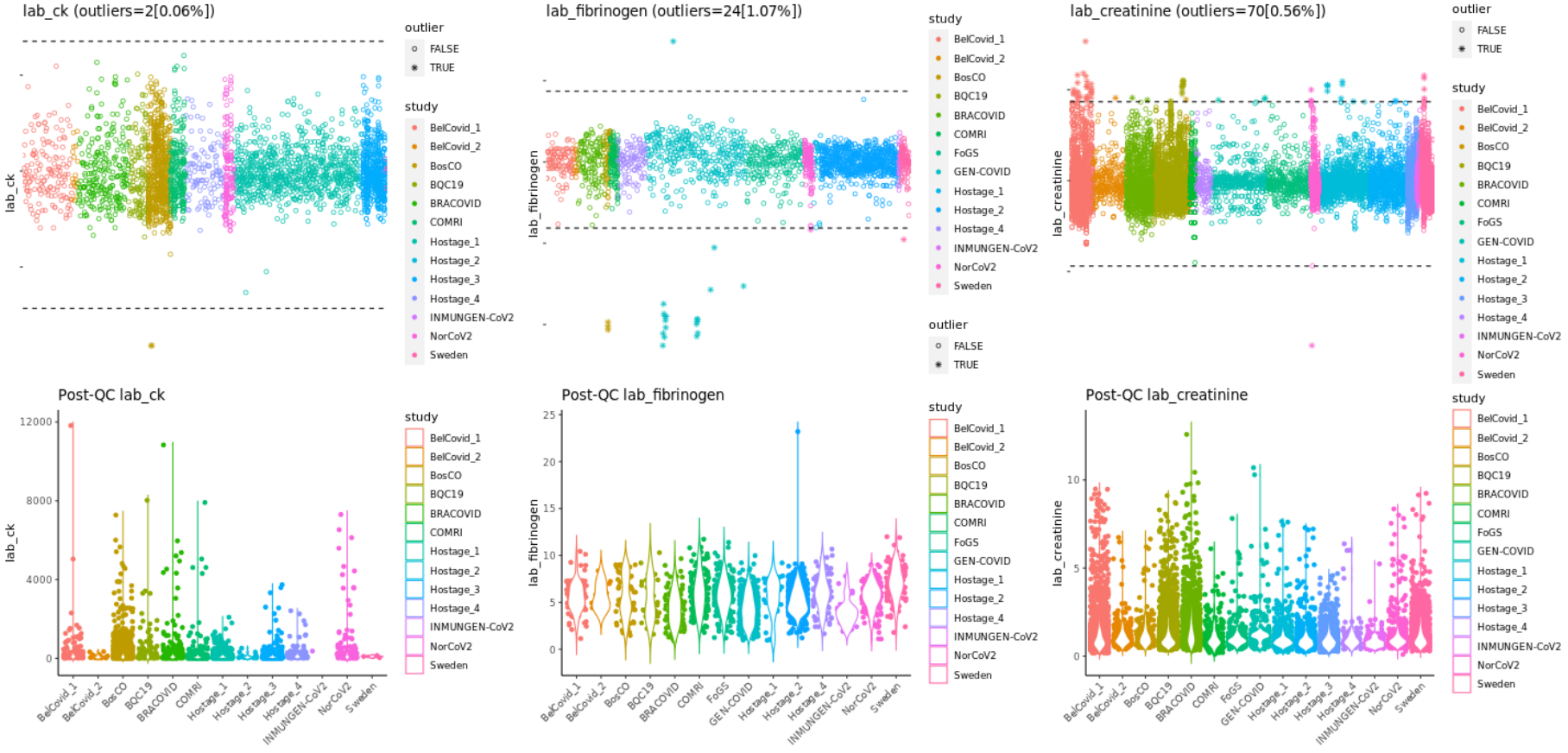

**Association with laboratory values**

From all the recorded values of a parameter per individual, we selected the highest/lowest value within -2 to 30 days from the COVID-19 diagnosis date, when the measurements at multiple time-points were available. We used the highest one for white blood cell counts^23^, neutrophil counts, monocyte counts, platelet counts, C-reactive protein, Troponin T, aspartate aminotransferase, alanine aminotransferase, total bilirubin, lactate dehydrogenase, γ-glutamyl transpeptidase, alkaline phosphatase, D-dimer, interleukin 6^20^, ferritin, fibrinogen^21^, procalcitonin, creatine kinase, and creatinine, whereas we used the lowest one for lymphocyte counts. We restricted this analysis to individuals of European descent. We imputed zero values (not detected in the biochemistry systems) to the lowest measurement amongst all individuals divided by two, and then natural log transformed the values, followed by regressing out the participating studies. The values were standardized for the coefficients to be comparable. Logistic regressions were fitted to assess the associations between the laboratory values and the severity phenotype, adjusted for age and sex as fixed effects and groups indicating participating studies as random effects. We selected death or severe respiratory failure as a severity outcome of COVID-19. Mixed linear regression was performed to assess the associations between chromosome 3 genetic risk allele carrier status and the laboratory values, adjusted for age, sex, genetic PCs 1 to 5 as fixed effects and groups indicating participating studies as random effects (see above) using “lme4 v1.1-25” R package. The results are described in **Supplementary Figure 3** and **Supplementary Table 2**.

###### Supplementary Figure 3: Bivariate plot to compare the coefficients of regressions between laboratory values and death or severe respiratory failure *vs* between rs10490770 risk allele carrier status and the laboratory values.

(x-axis) Coefficients of the logistic regressions between standardized laboratory values and death or severe respiratory failure. (y-axis) Coefficients of the linear regressions between rs10490770 risk allele carrier status and standardized laboratory values. Dots represent the point estimates for the coefficients, whereas lines represent the 95% confidence intervals. The dots and lines are solid when the p value < 0.05 for the association between rs10490770 risk allele carrier status and standardized laboratory values. WBC: white blood cell counts, Neut: neutrophil counts, Mono: monocyte counts, Plt: platelet counts, CRP: C-reactive protein, Trop-T: Troponin T, AST: aspartate aminotransferase, ALT: alanine aminotransferase, T-bil: total bilirubin, LDH: lactate dehydrogenase, GGT: γ-glutamyl transpeptidase, ALP: alkaline phosphatase, IL-6: interleukin 6, PCT: procalcitonin, CK: creatine kinase, Cre: creatinine, Lymph: lymphocyte counts.

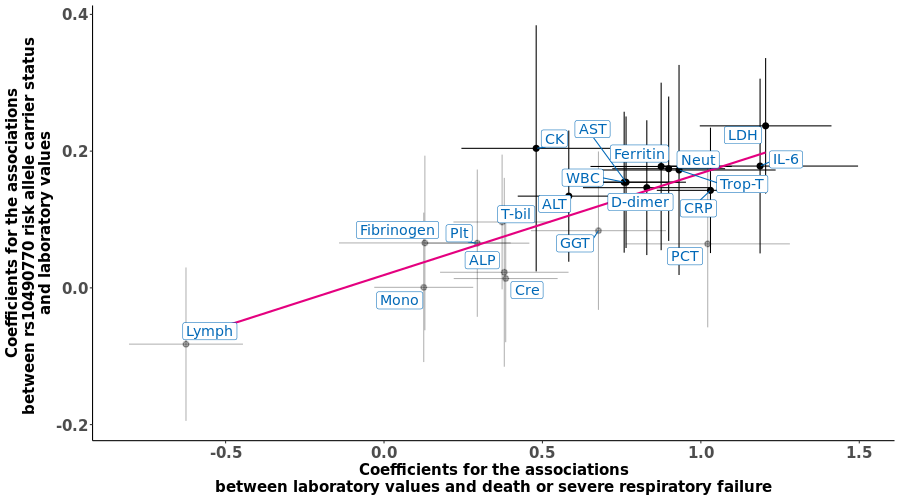

#### **Meta analyses**

As secondary analyses, we meta-analyzed the results with non-European ancestries and two external cohorts for which we did not have access to individual-level data; FinnGen and Columbia University COVID-19 Biobank (CUB). This resulted in a total study population of 14,625 individuals with COVID-19. An inverse-variance weighted meta-analyses were performed under a fixed effect and random effects models using the “meta v4·16-1” R package when the appropriate phenotypes were available and case counts, control counts, and the rs10490770 risk allele carrier counts were larger than ten in each cohort.

##### **Meta analyses of associations with mortality**

Meta-analyses across participants of European, South Asian and Admixed American descent on COVID-19 mortality were performed using an inverse-variance weighted models using the log-hazard ratio (HR) and its standard error as input. These provided similar estimates to the main analyses (HR for all-cause mortality 1.4, 95%CI 1.2–1.6, and HR for COVID-19 related death 1.5, 95%CI: 1.3–1.8, **Supplementary Figure 4**).

###### Supplementary Figure 4: Meta-analyses of associations with mortality

EUR: European, SAS: South Asian, AMR: Admixed-American, AFR: African, EAS: East Asian, HR: hazard ratio, 95%-CI: 95% confidence interval
(A) All-cause mortality (B) COVID-19 related mortality

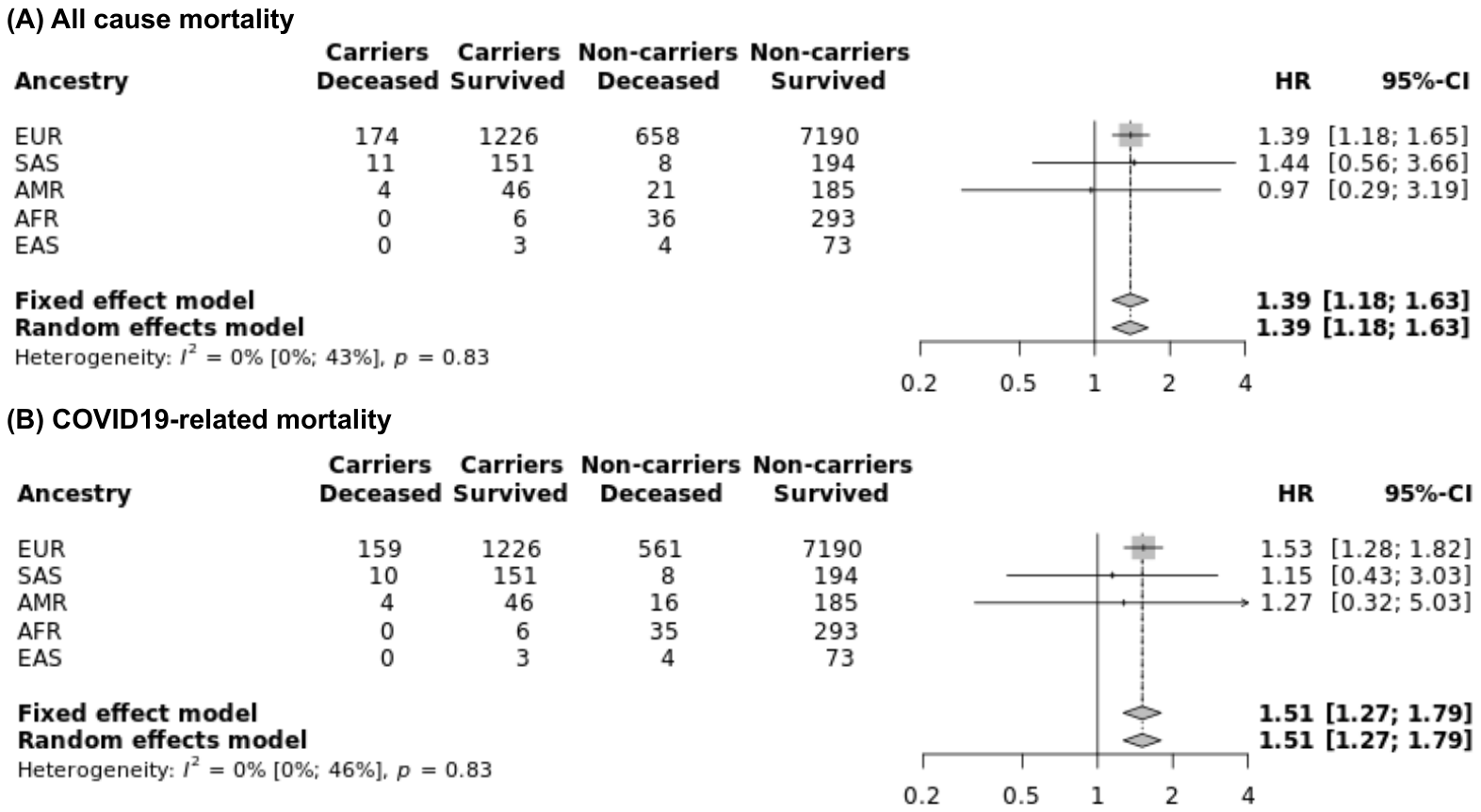

##### **Meta analyses of associations with COVID-19 severity**

Meta-analyses across participants of European, South Asian and Admixed American descent from COVID-19 HGI cohorts (those with individual level data) and one external cohort (FinnGen) on COVID-19 severity were performed using an inverse-variance weighted models using the log-odds ratio (OR) and its standard error as input. These provided similar estimates to the main analyses. Risk allele carrier status at rs10490770 was significantly associated with hospitalization (OR 1.5, 95%CI 1.3-1.7, p=8.5x10^-10^), ICU admission (OR 2.2, 95%CI 1.5-3.2, p=1.1x10^-4^ in random-effects model and OR 2.3, 95%CI 1.9-3.0, p=4.9x10^-13^ in a fixed-effect model) and death or severe respiratory failure (OR 1.7, 95%CI 1.5-2.0, p=2.7x10^-11^) (**Supplementary Figure 5**). The ancestries and cohorts were included when the appropriate phenotypes were available and case counts, control counts and the minor allele counts were larger than ten.

###### Supplementary Figure 5: Meta-analyses of associations with COVID-19 severity

COVID19-HGI-EUR: Individuals of European ancestry in COVID-19 HGI, COVID19-HGI-SAS: Individuals of South Asian ancestry in COVID-19 HGI, COVID19-HGI-AMR: Individuals of Admixed-American ancestry in COVID-19 HGI, COVID19-HGI-AFR: Individuals of African ancestry in COVID-19 HGI, COVID19-HGI-EAS: Individuals of East Asian ancestry in COVID-19 HGI, OR: odds ratio, 95%-CI: 95% confidence interval
(A) Hospitalization (B) ICU admission (C) Death or severe respiratory failure

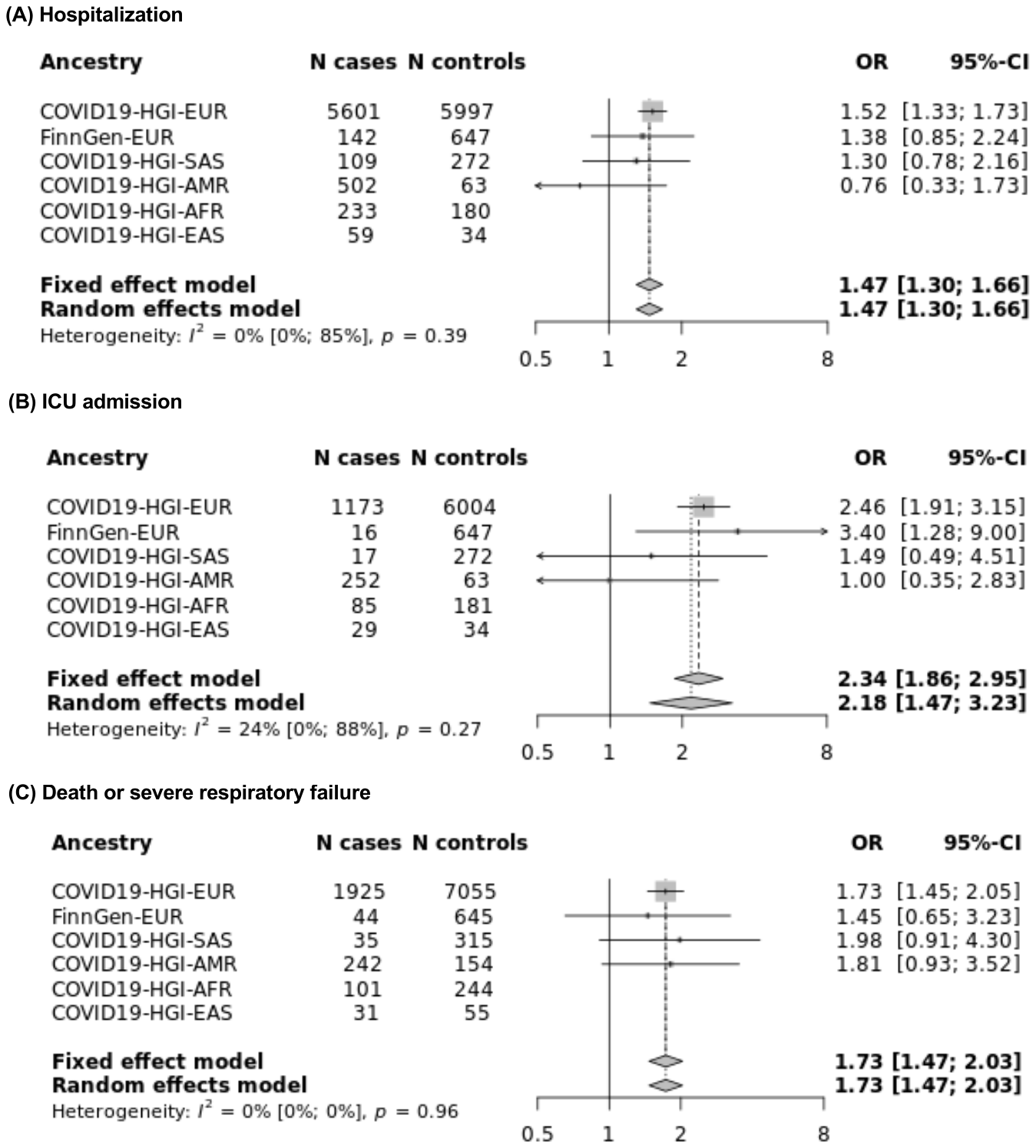

##### **Meta analyses of the associations with COVID-19 complications**

Meta-analyses across participants of European, South Asian and Admixed American descent from COVID-19 HGI cohorts (those with individual level data) and one external cohort (FinnGen) on COVID-19 severity were performed using an inverse-variance weighted models using the log-odds ratio (OR) and its standard error as input. These provided similar estimates to the main analyses. Risk allele carrier status at rs10490770 was significantly associated with severe respiratory failure (OR 2.0, 95%CI 1.6-2.4, p=2.6x10^-11^ both under fixed and random-effects model) and hepatic injury (OR 1.5, 95%CI 1.1-1.9, p=3.3x10^-3^ in a fixed-effect model and OR 1.5, 95%CI 1.1-2.0, p=7.6x10^-3^ in random-effects model). A fixed effect model provided a similar effect for venous thromboembolism (OR 1.6, 95%CI 1.2-2.2, p=2.8x10^-3^) but not in random-effects model (OR 1.2, 95%CI 0.44-3.2, p=0.74) (**Supplementary Figure 6**). The ancestries and cohorts were included when the appropriate phenotypes were available and case counts, control counts and the minor allele counts were larger than ten.

###### Supplementary Figure 6: Meta-analyses of the associations with COVID-19 complications

COVID19-HGI-EUR: Individuals of European ancestry in COVID-19 HGI, COVID19-HGI-SAS: Individuals of South Asian ancestry in COVID-19 HGI, COVID19-HGI-AMR: Individuals of Admixed-American ancestry in COVID-19 HGI, COVID19-HGI-AFR: Individuals of African ancestry in COVID-19 HGI, COVID19-HGI-EAS: Individuals of East Asian ancestry in COVID-19 HGI FinnGen-EUR: Individuals of European ancestry in FinnGen. OR: odds ratio, 95%-CI: 95% confidence interval.
(A) Severe respiratory failure (B) Hepatic injury (C) Cardiovascular complications (D) Kidney injury (E) Venous thromboembolism

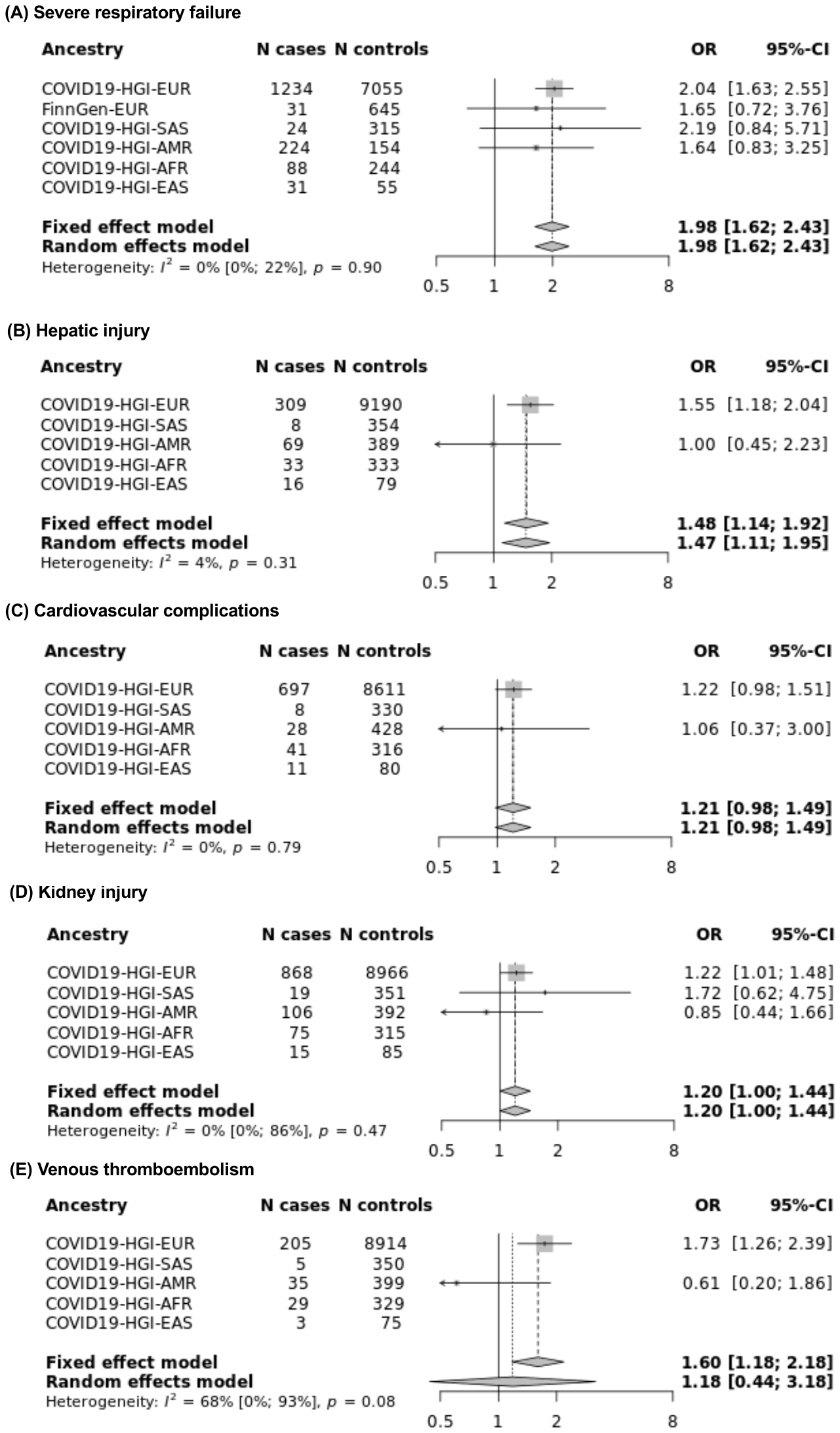

##### **Meta analyses of linear regressions between risk allele carrier status and age at diagnosis of severe COVID-19.**

Meta-analyses across participants of European, South Asian, Admixed American, and African descent from COVID-19 HGI cohorts (those with individual level data) and one external cohort (FinnGen, CUB) on COVID-19 severity were performed using an inverse-variance weighted models using the coefficient and its standard error as input. Amongst patients who admitted to ICU, risk allele carriers were on average 2.1 years younger than non-carriers (95%CI 0.53-3.6, p=8.3x10^-3^ under both fixed and random effects model). Similarly, amongst patients who died or experienced severe respiratory failure, risk allele carriers were on average 1.9 years younger than non-carriers (95%CI 0.79-3.1, p=8.6x10^-4^ under both fixed and random effects model). However, similar trend was not confirmed amongst hospitalized patients. (Mean difference -0.67, 95%CI -1.5 0.13, p=0.099 under fixed-effects model) (**Supplementary Figure 7**). The ancestries and cohorts were included when the appropriate phenotypes were available and case counts were larger than ten and the minor allele counts amongst cases were larger than three.

###### Supplementary Figure 7: Meta analyses of linear regressions between risk allele carrier status and age at diagnosis of severe COVID-19.

COVID19-HGI-EUR: Individuals of European ancestry in COVID-19 HGI, COVID19-HGI-SAS: Individuals of South Asian ancestry in COVID-19 HGI, COVID19-HGI-AMR: Individuals of Admixed-American ancestry in COVID-19 HGI, COVID19-HGI-AFR: Individuals of African ancestry in COVID-19 HGI, COVID19-HGI-EAS: Individuals of East Asian ancestry in COVID-19 HGI FinnGen-EUR: Individuals of European ancestry in FinnGen. CUB-EUR: Individuals of European ancestry in CUB. CUB-AMR: Individuals of Admixed-American ancestry in CUB. CUB-AFR: Individuals of African ancestry in CUB. Coefficients: coefficients for linear associations between age and carrier status, 95%-CI: 95% confidence interval.
(A) Hospitalization (B) ICU admission (C) Death or severe respiratory failure

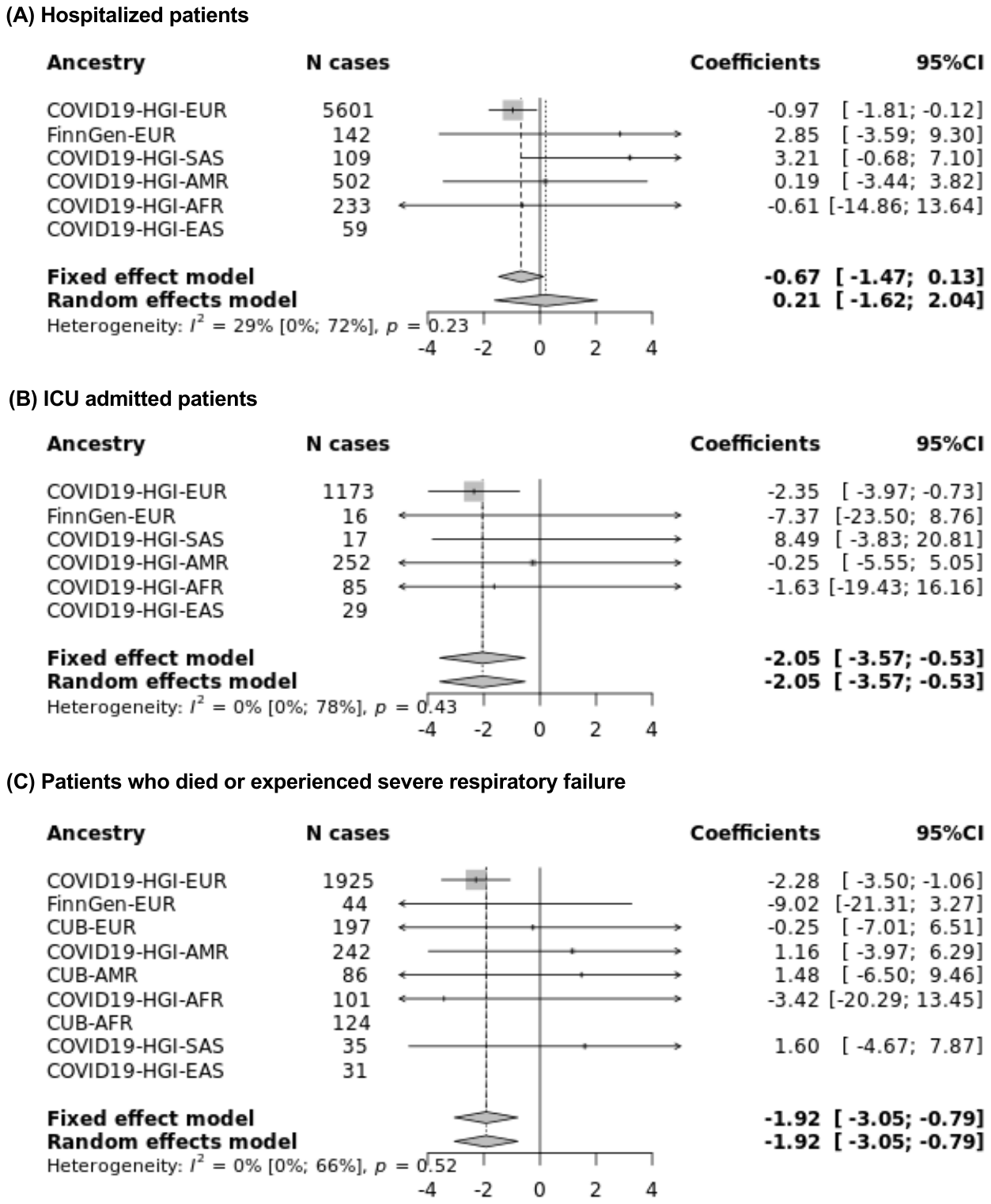

### References

1	COVID-19 Host Genetics Analysis Plan v1.1 - Google Docs. https://docs.google.com/document/d/16ethjgi4MzlQeO0KAW_yDYyUHdB9kKbtfuGW4XYVKQg/edit#heading=h.yvzvdf3jx6u9 (accessed Feb 7, 2021).

2	(No Title). https://www.thermofisher.com/order/catalog/product/902981#/902981 (accessed Jan 22, 2021).

3	Auton A, Abecasis GR, Altshuler DM, *et al.* A global reference for human genetic variation. Nature. 2015; **526**: 68–74.

4	The NHLBI Trans-Omics for Precision Medicine (TOPMed) Whole Genome Sequencing Program. BRAVO variant browser: University of Michigan and NHLBI. 2018. https://bravo.sph.umich.edu/freeze5/hg38/ (accessed Aug 28, 2020).

5	Taliun D, Harris DN, Kessler MD, *et al.* Sequencing of 53,831 diverse genomes from the NHLBI TOPMed Program. *bioRxiv* 2019; **2**: 563866.

6	Das S, Forer L, Schönherr S, *et al.* Next-generation genotype imputation service and methods. *Nat Genet* 2016; **48**: 1284–7.

7	Bycroft C, Freeman C, Petkova D, *et al.* The UK Biobank resource with deep phenotyping and genomic data. *Nature* 2018; **562**: 203–9.

8	Genotyping and quality control of UK Biobank, a large-scale, extensively phenotyped prospective resource. http://biobank.ctsu.ox.ac.uk. (accessed Feb 18, 2021).

9	Shrine N, Guyatt AL, Erzurumluoglu AM, *et al.* New genetic signals for lung function highlight pathways and chronic obstructive pulmonary disease associations across multiple ancestries. *Nat Genet* 2019; **51**: 481–93.

10	Karczewski KJ, Francioli LC, Tiao G, *et al.* The mutational constraint spectrum quantified from variation in 141,456 humans. *Nature* 2020; **581**: 434–43.

11	Ellinghaus D, Degenhardt F, Bujanda L, *et al.* Genomewide Association Study of Severe Covid-19 with Respiratory Failure. *N Engl J Med* 2020; : NEJMoa2020283.

12	Purcell S, Neale B, Todd-Brown K, *et al.* PLINK: A tool set for whole-genome association and population-based linkage analyses. *Am J Hum Genet* 2007; **81**: 559–75.

13	Price AL, Weale ME, Patterson N, *et al.* Long-Range LD Can Confound Genome Scans in Admixed Populations. Am. J. Hum. Genet. 2008; **83**: 132–5.

14	Abraham G, Qiu Y, Inouye M, Stegle O. Genetics and population analysis FlashPCA2: principal component analysis of Biobank-scale genotype datasets. DOI:10.1093/bioinformatics/btx299.

15	Pärn K, Fontarnau JN, Isokallio MA, *et al.* Genotyping chip data lift-over to reference genome build GRCh38/hg38 V.1. 2018; published online April 13. DOI:10.17504/protocols.io.nqtddwn.

16	Pärn K, Isokallio MA, Fontarnau JN, Palotie A, Ripatti S. Genotype imputation workflow v3.0 V.1. 2018; published online May 10. DOI:10.17504/protocols.io.nmndc5e.

17	Altshuler DM, Gibbs RA, Peltonen L, *et al.* Integrating common and rare genetic variation in diverse human populations. *Nature* 2010; **467**: 52–8.

18	Hail Team. Hail 0.2. https://github.com/hail-is/hail. .

19	Horita N, Kaneko T. Genetic model selection for a case-control study and a meta-analysis. *Meta Gene* 2015; **5**: 1–8.

20	Del Valle DM, Kim-Schulze S, Huang H-H, *et al.* An inflammatory cytokine signature predicts COVID-19 severity and survival. *Nat Med* 2020; : 1–8.

21	Merrill JT, Erkan D, Winakur J, James JA. Emerging evidence of a COVID-19 thrombotic syndrome has treatment implications. Nat. Rev. Rheumatol. 2020; **16**: 581–9.

22	Higuera-de la Tijera F, Servín-Caamaño A, Reyes-Herrera D, *et al.* Impact of liver enzymes on SARS-CoV-2 infection and the severity of clinical course of COVID-19. *Liver Res* 2021; published online Jan 12. DOI:10.1016/j.livres.2021.01.001.

23	Vafadar Moradi E, Teimouri A, Rezaee R, *et al.* Increased age, neutrophil-to-lymphocyte ratio (NLR) and white blood cells count are associated with higher COVID-19 mortality. *Am J Emerg Med* 2021; **40**: 11–4.

24	Yan L, Zhang H-T, Goncalves J, *et al.* An interpretable mortality prediction model for COVID-19 patients. *Nat Mach Intell* 2020; **2**: 283–8.
